# Cancer Stem Cell–Associated Marker Expression in Chemotherapy-Treated Wilms Tumour

**DOI:** 10.64898/2026.03.17.26348535

**Authors:** Masoumeh Mousavinejad, Lisa Howell, Patricia Murray, Ed Cheesman, Barry Pizer, Paul D. Losty, Srinivas Annavarapu, Rajeev Shukla, Bettina Wilm

## Abstract

**Background:** Wilms tumour (WT) relapse occurs more frequently in patients with blastemal-type WTs. The presence of cancer stem cells (CSCs) is linked to tumour survival and relapse, and CSCs may be found in greater numbers in blastemal cell foci. CSC-associated phenotypes have been described in untreated WT, but their persistence, organisation and relevance after neoadjuvant chemotherapy is unknown.

**Methods:** We analysed 23 formalin-fixed paraffin-embedded blocks from 18 chemotherapy-treated patients where WTs were enriched for viable blastema, using human fetal kidney as developmental control. Immunohistochemistry and -fluorescence analysis determined progenitor (PAX2, SIX2, CITED1) and CSC-associated (NCAM, ALDH1, CD133) marker expression. We qualitatively and semi-quantitatively evaluated spatial expression patterns and co-localisation across tumour compartments.

**Results:** PAX2 and SIX2 were co-expressed in blastema in most cases (15/18), with PAX2 expression higher at the periphery of blastemal foci and SIX2 expression found uniformly in central aspects. CITED1 expression was also associated with SIX2 in blastema tissues (14/18). NCAM was blastema-enriched (15/18) with higher central intensity, frequently adjacent to PAX2-expressing peripheral zones. ALDH1 expression was present across blastema and epithelium while NCAM-, ALDH1-double-positive cells were rarely observed (4/18). CD133 expression was less commonly seen (2/18), localising near epithelial/nephrogenic structures.

**Conclusions:** After neoadjuvant chemotherapy, WT blastema retained overlapping but non-identical progenitor/CSC-associated marker landscapes with reproducible peripheral–centre gradients. These spatial arrangements suggest a blastemal niche for CSCs that may sustain a therapy-resistant state. Our analysis provides the foundation for future functional validation and molecular profiling to define key lineage relationships and therapeutic vulnerabilities in post-chemotherapy WT. [250/250 words]

## Introduction

Wilms tumour (WT) is the most common kidney cancer in children, with up to 80 new patients diagnosed annually in the UK. WT arises from kidney progenitor tissues during embryonic development of the kidney (Li et al., 2021). In the UK, curative therapies involve pre-operative vincristine and actinomycin D +/- doxorubicin treatment based on the International Society of Pediatric Oncology Renal Tumour Study Group (SIOP-RTSG) strategy. Following scheduled nephrectomy, post-operative chemotherapy and radiotherapy is based on post-tumour nephrectomy staging. This strategy achieves an excellent overall 5-year survival rate of over 85% (Fawkner-Corbett et al., 2014). The US-based Children’s Oncology Group (COG), sees similar successful survival rates by performing an initial ‘up front’ nephrectomy, followed by staging-directed therapy including chemotherapy and/or radiotherapy (Groenendijk et al., 2021). Independent of strategy approach, success rates are similar (Vujanic et al., 2022). However, despite a good overall success rate, around 10-15% of patients with WT will experience relapse. The most common sites of relapse are the lungs followed by abdominal sites and liver. Whilst relapse treatments are often successful, there are higher risk groups for whom outcome is poor and additional treatments are linked with associated morbidity (Losty, 2016; Pasqualini et al., 2020; van den Heuvel-Eibrink et al., 2017).

WTs typically contain 3 different histological cell types (triphasic) recognised as stromal, epithelial and blastemal. There is a considerable amount of variability in the proportional composition of these cell types. Tumours with predominance of blastemal cells are termed ‘blastemal type’ tumours. The risk of relapse is significantly higher in patients with these high-risk tumour types (Ortiz et al., 2023; Pasqualini et al., 2020; van den Heuvel-Eibrink et al., 2015). The blastema in WT most closely resembles the metanephric mesenchyme in the developing kidney and is composed of progenitor cells that give rise to the majority of epithelial and stromal kidney cells and tissues. It is important to recognise that neo adjuvant chemotherapy has an impact on the resulting composition of cell types and therefore the SIOP and COG categorisation of tumour types are not directly comparable (Popov et al., 2016; Vujanic et al., 2022). It is now well reported that post chemotherapy residual blastema are associated with a poorer outcome and higher risk of relapse (Spreafico et al., 2021). Work to quantify the relevant volumes which infer a significant worse outcome is underway with the hope to further stratify patients on this basis.

In WT tissue, blastema is a heterogeneously differentiating cell population that follows an organotypical mesenchymal to epithelial transition (MET) process as observed during kidney development (Shukrun et al., 2014). SIX2, PAX2 and CITED1 are the main transcription factors that have been extensively studied in the context of WT, as they are considered markers of the blastemal component (Petrosyan et al., 2023; Royer-Pokora et al., 2023; Sehic et al., 2014). During normal development of nephrons, SIX2, PAX2 and CITED1 expression is tightly regulated playing a crucial role in maintaining the nephron progenitor population and balancing self-renewal and differentiation processes (Dickinson et al., 2022; Khoshdel Rad et al., 2020). The expression of these three key markers is localised in nephrogenic zones, indicating their involvement in nephron formation, but is downregulated and ultimately lost when the respective cells finally differentiate into mature nephron structures (Li et al., 2021; Pode-Shakked et al., 2013).

A current concept that may be key to understanding WT biology, suggests that WT blastema harbour a remaining pool of progenitor cells, which are the source of cancer stem cells (CSCs) (Brok et al., 2018). CSCs are a subset of cells within tumours possessing properties of self-renewal and multilineage differentiation abilities. CSCs contribute to tumour recurrence and treatment resistance in cancer patients, frequently rendering cancer incurable (Marzagalli et al., 2021). Extensive research has been conducted to characterise the molecular profile of CSCs in various cancers and to explore their response to pharmacological targeting as therapeutic interventions to block tumour growth and disease relapse (Philchenkov and Dubrovska, 2024). Consequently, in relapsing WT, it may be blastemal-type cells with stem or progenitor cell characteristics that contribute to driving relapse (Pode-Shakked et al., 2013; Shukrun et al., 2014; Spreafico et al., 2009).

Increasing interest in CSCs and their pivotal roles in cancer biology has led to the identification of CSC in WT, based on a range of markers, including CD133 (Jafarian et al., 2020; Mehrazma et al., 2013; Singh et al., 2023), NCAM and its co-expression with ALDH1 (Pode-Shakked et al., 2009; Pode-Shakked et al., 2013; Raved et al., 2019) or SIX2 (Pierce et al., 2014), and co-expression of SIX2 and CITED1 (Murphy et al., 2014; Murphy et al., 2012; Petrosyan et al., 2023). Experimental studies using WT-derived xenografts in mice have demonstrated the existence of ‘cancer initiating cells’ or CSCs which co-expressed NCAM and ALDH1 (Pode-Shakked et al., 2013). Furthermore, NCAM-, ALDH1-double positive CSC in blastemal WT were found to correlate with poor clinical outcome and disease progression (Raved et al., 2019).

Whilst these studies have determined the identities and functional roles of CSCs within WT, they are based on the analysis of naïve, untreated WT samples. To the best of our knowledge, there have been no research studies published which specifically characterise CSCs and kidney progenitor cells associated with blastemal cells in tumours treated with neoadjuvant chemotherapy, as per SIOP strategy. There is therefore a lack of understanding of the impact of chemotherapy on blastemal associated-CSCs and the tumour microenvironment. This may explain their potential to lead to disease progression, metastasis formation and poor clinical outcomes.

To address this knowledge gap, we performed an exploratory and descriptive assessment of archived WT nephrectomy specimens collected post neoadjuvant chemotherapy. We determined the co-expression of a range of CSC-associated (NCAM, ALDH1 and CD133) and kidney progenitor (PAX2, SIX2, CITED1) markers. We also described the spatial arrangement of these cells within tumours. Despite expected case-to-case variations in WT specimens, our results demonstrate intense co-expression of kidney progenitor cell- and CSC-associated markers within blastema tissues, but also differential expression of some of these markers. While the sample size was small, these findings may offer insights into the molecular underpinnings of WT persistence and relapse.

## Methods

### Sample collection and ethics statement

Specimens were collected from 18 WT index patients where parents/guardians had provided informed consent to the UK VIVO Biobank (project number 23-VIVO-10), and/or the UK IMPORT study for renal tumours of childhood (REC reference 12/LO/0101). Foetal human kidney (hFK) specimens from elective terminations at 22–24 weeks of gestational age, were obtained after full informed consent from parents (REC reference 25/PR/0493). Institutional sponsorship was granted for this study and full ethics approval obtained from the relevant research committees. The study followed all applicable regulations and ethical standards for the use of human tissue in biomedical research and adhered to the principles of the Declaration of Helsinki.

A total of 23 WT paraffin-embedded tissue blocks (from 18 patients) as well as one human foetal kidney tissue specimen (stage 22 weeks) were analysed. These samples were reviewed by expert histopathologists and selected based on pre-treated turmours found to have triphasic elements with viable blastemal regions.

All WT and foetal kidney cases underwent immunohistochemistry (IHC) testing to evaluate the expression of PAX2, SIX2, CITED1, NCAM, ALDH1, and CD133. For co-expression analyses, a subset of samples was randomly selected to undergo dual IHC staining for PAX2/SIX2 and SIX2/CITED1 combinations allowing for the assessment of overlapping progenitor marker expression within blastemal regions. In cases showing apparent co-expression of NCAM and ALDH1 by IHC, dual immunofluorescence (IF) staining was subsequently performed to confirm true co-localisation of these markers within the same cell populations.

Specimens were fully anonymised before investigative analysis, and the research team remained blinded to clinical course and patient outcome data. For completeness, in Supplementary Table 1 we present the relevant clinical disease staging, treatment and outcomes.

### Immunohistochemical (IHC) staining

Immunohistochemical staining was performed on formalin-fixed, paraffin-embedded (FFPE) Wilms tumour tissue sections. Sections cut at 5 μm were dried overnight at 37 °C, de-waxed in fresh xylene and rehydrated through a graded ethanol series (100%, 95%, 70%) before rinsing in distilled water. Antigen retrieval was performed using heat-induced epitope retrieval (HIER) in citrate buffer (pH 6.0) for 20 minutes at near-boiling temperature. Subsequently, cooled slides were quenched with 3% hydrogen peroxide for 10 minutes followed by treatment with blocking solution (5% normal serum matched to the secondary antibody host species).

Primary antibodies were applied at optimised dilutions and incubated overnight at 4 °C followed by washes in phosphate-buffered saline (PBS) and incubation for 30 minutes at room temperature with species-specific biotinylated or HRP-conjugated secondary antibodies. Antibody binding was detected using 3,3′-diaminobenzidine (DAB) substrate solution (Vector Laboratories, UK) and sections counterstained with haematoxylin, dehydrated in ethanol, cleared in xylene, and mounted with DPX mounting medium.

A complete list of all primary antibodies, including host species, source, catalogue numbers, and working dilutions, is provided in Supplementary Table 2.

### Immunofluorescence (IF) staining

Immunofluorescence staining was performed on 5 μm formalin-fixed, paraffin-embedded (FFPE) tissue sections and processed through HIER antigen retrieval. Subsequently, tissue sections were permeabilised with 0.1% (v/v) Triton X-100 in PBS for 10 minutes and blocked with 1% (w/v) BSA in PBS for 30 minutes at room temperature. Primary antibodies diluted in blocking buffer were applied overnight at 4 °C in a humidified chamber. After washing in PBS, sections were incubated with fluorophore-conjugated secondary antibodies for 1 hour at room temperature, counterstained with DAPI, and mounted with anti-fade fluorescent mounting medium (Dako, Agilent).

A full list of primary and secondary antibodies, including host species, fluorophore conjugates, catalogue numbers, and working dilutions, is provided in Supplementary Table 2.

### Image analysis

Stained slides after IHC were digitally scanned at high resolution using a Leica Aperio CS Slide Scanner. Image analysis and quantification were performed using QuPath, Fiji (ImageJ), and ImageScope. H-scores for each marker were generated in QuPath using automated thresholding parameters established from normal fetal kidney tissue as control. Scores were grouped as Low (0–100), Moderate (101–200), and High (201–300), based on these control-derived thresholds. All scoring and subsequent validation were conducted by a single investigator blinded to the clinical and pathological data. For downstream analyses, marker expression was manually binarised (1 = expression present, 0 = no expression), with Moderate and High scores considered as expression (Supplementary Table 3 for detailed information). The decision to binarise was based on the limited sample size (n = 18), which reduced statistical power for quantitative analyses.

Fluorescent images were acquired using a 3i spinning disk confocal microscope (CSU-X1, Intelligent Imaging Innovations). Multiple *z*-stacks were collected where appropriate to capture three-dimensional distribution of signals. Images were exported as TIFF files and processed using Fiji (ImageJ) software. Pre-processing included background subtraction, intensity normalisation and channel separation.

Quantification of marker expression was performed using region of interest (ROI)-based measurements, and co-localisation analysis was assessed using Pearson’s correlation coefficients. The Radial Profile plug-in was employed to analyse spatial distribution of fluorescence intensity relative to defined cellular landmarks. Where applicable, cell counts, and fluorescence intensity values were normalised to DAPI-positive nuclei to account for differences in tissue density.

### Statistical analysis

For IHC, pairwise relationships between markers were quantified using GraphPad Prism to generate a similarity matrix, which was visualised as a Circos-style network.

For IF, partial co-localisation of markers was assessed using Pearson’s correlation coefficient (PCC, r) with the Coloc2 plugin in Fiji (ImageJ). Coefficients were calculated across five distinct regions of interest (ROIs) per case and averaged to obtain representative values, with r ranging from –1 (perfect negative correlation) to +1 (perfect positive correlation).

## Results

### PAX2 and SIX2 co-expression

We identified and located kidney progenitor cells in sections of 18 WT index cases (Table 1) using antibodies against the established kidney progenitor markers PAX2 and SIX2 (Davis et al., 2011; Eccles et al., 1992; Pleniceanu et al., 2010). In human foetal kidney (hFK) sections, PAX2, a key regulator of nephrogenesis, was expressed in cells undergoing mesenchymal-to-epithelial transition (MET). It labelled the cap mesenchyme and its early derivatives, including comma and S-shaped bodies in the peripheral nephrogenic zones (NZ). PAX2 expression was also found in immature tubules of the kidney medulla (M) (Supplementary Figure 1A, B). SIX2, a direct regulator of the self-renewal of nephric progenitor cells (Kobayashi et al., 2008), was expressed in PAX2-positive domains but was absent from immature tubules in adjacent hFK sections (Supplementary Figure 1C, D).

**Table 1:** Distribution of markers used in the study for WT patients. Blastemal (B), Epithelial (E), and Stromal (S) components.

| Marker | PAX2 |  |  | SIX2 |  |  | CITED1 | NCAM | ALDH1 | NCAM<sup>+</sup><br>ALDH1<sup>+</sup> | CD133 | Blastemal co-expression |
| --- | --- | --- | --- | --- | --- | --- | --- | --- | --- | --- | --- | --- |
| Case | B | E | S | B | E | S |  |  |  |  |  |  |
| 1 | ✓ | ✓ | ✗ | ✓ | ✗ | ✗ | ✓ | ✗ | ✓ | ✗ | ✗ | PAX2/ SIX2/ CITED1 |
| 2 | ✓ | ✓ | ✓ | ✗ | ✗ | ✗ | ✓ | ✗ | ✓ | ✗ | ✗ | PAX2/ CITED1 |
| 3 | ✓ | ✓ | ✗ | ✓ | ✗ | ✗ | ✓ | ✓ | ✓ | ✗ | ✗ | PAX2/ SIX2/ CITED1/ NCAM |
| 4 | ✓ | ✓ | ✗ | ✓ | ✗ | ✗ | ✓ | ✓ | ✓ | ✓ | ✗ | PAX2/ SIX2/ CITED1/ NCAM<br>NCAM/ ALDH1 |
| 5 | ✓ | ✓ | ✗ | ✓ | ✗ | ✓ | ✓ | ✓ | ✓ | ✗ | ✗ | PAX2/ SIX2/ CITED1/ NCAM |
| 6 | ✓ | ✓ | ✗ | ✓ | ✓ | ✗ | ✗ | ✓ | ✓ | ✗ | ✗ | PAX2/ SIX2/ NCAM |
| 7 | ✓ | ✓ | ✗ | ✓ | ✗ | ✗ | ✓ | ✓ | ✓ | ✓ | ✓ | PAX2 /SIX2/ CITED1/ NCAM<br>NCAM/ ALDH1 |
| 8 | ✓ | ✓ | ✗ | ✓ | ✗ | ✗ | ✓ | ✓ | ✗ | ✗ | ✗ | PAX2/ SIX2/ CITED1/ NCAM |
| 9 | ✗ | ✗ | ✗ | ✓ | ✓ | ✗ | ✓ | ✓ | ✓ | ✗ | ✗ | SIX2/ CITED1/ NCAM |
| 10 | ✓ | ✓ | ✗ | ✓ | ✗ | ✗ | ✗ | ✓ | ✓ | ✗ | ✗ | PAX2/ SIX2/ NCAM |
| 11 | ✓ | ✓ | ✗ | ✓ | ✗ | ✗ | ✓ | ✓ | ✓ | ✓ | ✗ | PAX2/ SIX2/ CITED1/ NCAM<br>NCAM/ ALDH1 |
| 12 | ✓ | ✓ | ✗ | ✓ | ✗ | ✗ | ✓ | ✓ | ✓ | ✗ | ✗ | PAX2/ SIX2/ CITED1/ NCAM |
| 13 | ✓ | ✓ | ✗ | ✓ | ✗ | ✗ | ✓ | ✓ | ✓ | ✗ | ✗ | PAX2/ SIX2/ CITED1/ NCAM |
| 14 | ✓ | ✗ | ✗ | ✓ | ✓ | ✗ | ✓ | ✓ | ✓ | ✗ | ✗ | PAX2/ SIX2/ CITED1/ NCAM |
| 15 | ✓ | ✓ | ✗ | ✓ | ✗ | ✗ | ✓ | ✓ | ✓ | ✗ | ✗ | PAX2/ SIX2/ CITED1/ NCAM |
| 16 | ✓ | ✓ | ✗ | ✓ | ✗ | ✗ | ✓ | ✓ | ✓ | ✗ | ✗ | PAX2/ SIX2/ CITED1/ NCAM |
| 17 | ✓ | ✓ | ✗ | ✓ | ✗ | ✗ | ✓ | ✓ | ✓ | ✓ | ✓ | PAX2/ SIX2/ CITED1/ NCAM<br>NCAM/ ALDH1 |
| 18 | ✗ | ✓ | ✗ | ✗ | ✗ | ✗ | ✗ | ✗ | ✓ | ✗ | ✗ | x |

Overall, PAX2 was expressed in 16 of 18 WT cases (Table 1, Supplementary Table 3, Supplementary Figure 5). To determine the distribution of the expression pattern between the different WT tissue components, blastema, epithelium and stroma, we performed more detailed analysis and found that in 15 out of 18 WT patients, PAX2 was expressed in both the blastema and epithelial components, whilst in one single WT case, it was also detected in the stromal compartment (Table 1, Supplementary Figure 5). Overall, SIX2 expression was detected in 16 of 18 WT cases (Table 1, Supplementary Table 3, Supplementary Figure 5), specifically in the blastemal component. SIX2 was additionally expressed in the epithelial component of 3 cases and in the stromal compartment in a further single patient (Table 1). Overlapping expression domains of PAX2 and SIX2 were observed in the blastema in 15 WT cases (Figure 1A-D, Table 1, Supplementary Figure 2). Pearson’s correlation analysis in WT1 tumour case 1, revealed high correlation with a mean of 0.65 (+/- 0.04 SD) for PAX2 and SIX2 co-expression in 5 selected regions of interest. In WT tumour case 6, intense nuclear PAX2 expression and weak to faint SIX2 expression was detected in the epithelial compartments, consistent with previously described expression patterns in naïve WT specimens (Davis et al., 2011; Murphy et al., 2012). Expression levels of PAX2 varied within the blastemal tissue regions, with more intense staining at the periphery and weaker signal at the centre, whereas SIX2 was more uniformly expressed throughout the blastema (Figure 2A, B). This pattern was consistently observed in all WT specimens analysed where PAX2 and SIX2 were co-expressed in the blastema.

**Figure 1.**
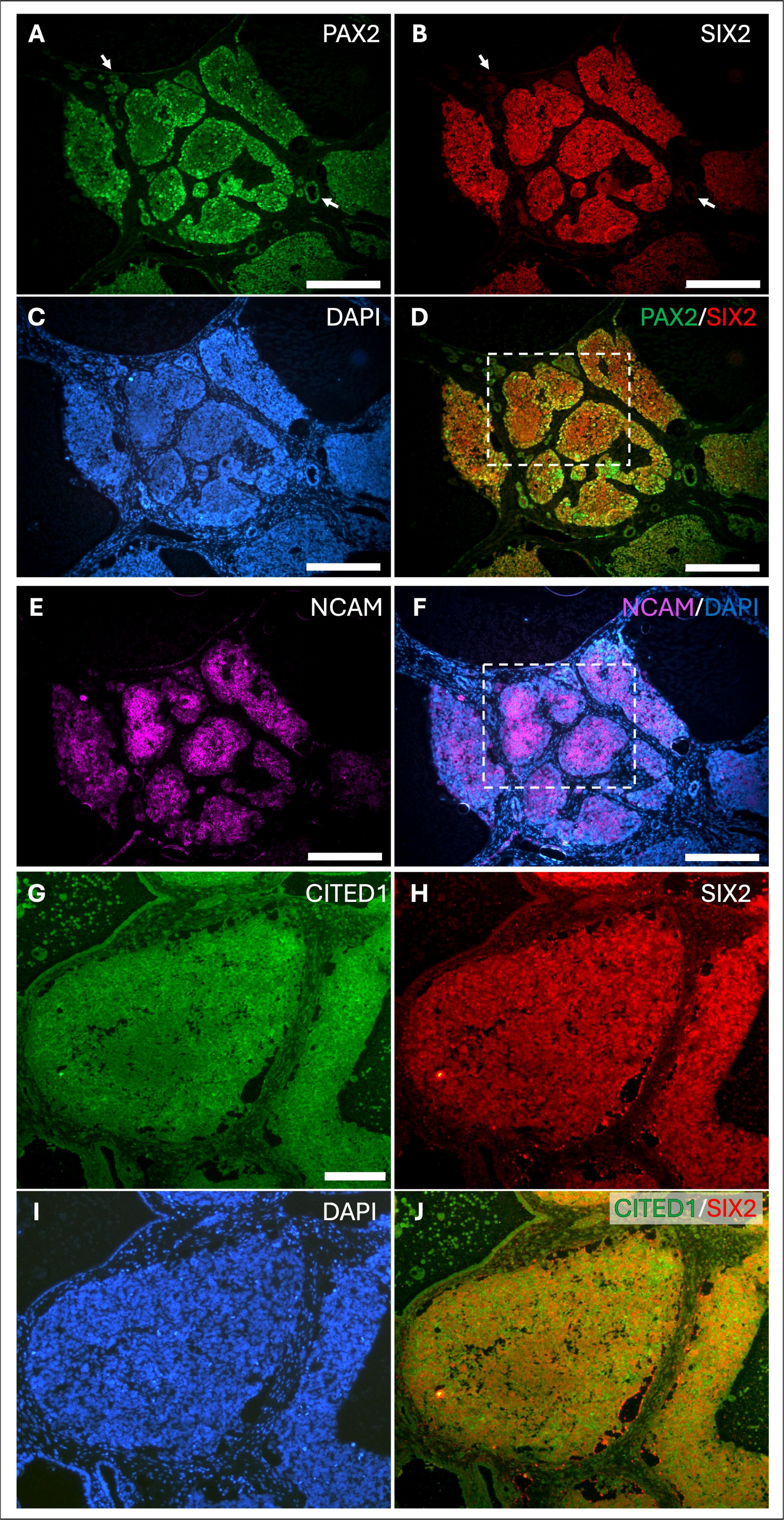
Co-expression of PAX2, SIX2, CITED1 and NCAM in WT. A-D. Double IF staining of SIX2 and PAX2 in a triphasic region in index WT case 3 showed immune-positivity of both markers in blastema cells while areas of epithelial differentiation were only positive for PAX2 (white arrows). White box in D outlines area shown in Figure 2A. **E, F.** IF staining of NCAM on a serial section. NCAM expression was most intense within blastema elements. White box in F outlines area shown in Figure 2C. **G-J.** Double IF staining of SIX2 and CITED1 in index WT case 3 showed a similar expression pattern in the blastema to that of SIX2 and PAX2. While SIX2 had nuclear localisation in blastemal cells, CITED1 was predominantly expressed in the cytosol. Scale bars are 100 µm (A-F), 50 µm (G-J).

**Figure 2.**
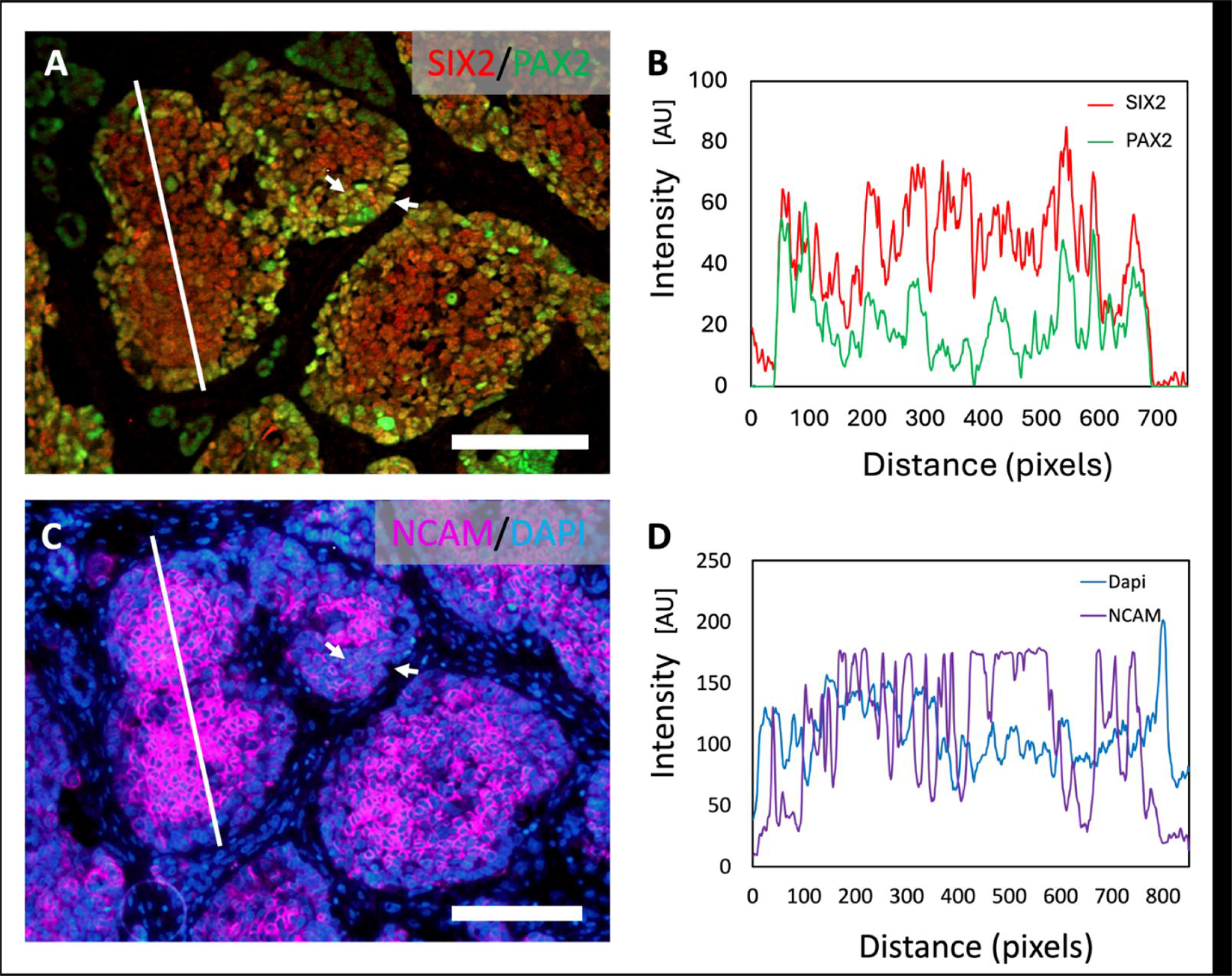
Analysis of heterogeneous expression of PAX2, SIX2 and NCAM in two serial sections of WT blastema tissue. A. High power magnification of area indicated in Figure 1D. Highest PAX2 intensity was localised at the edges of blastema (white arrows), while SIX2 was expressed more uniformly throughout the blastema in WT case 3. B. Quantification of signal intensities for PAX2 and SIX2 across the white line in A (blastema) reflects that PAX2 expression levels were lower within the blastema centre, while SIX2 was highly expressed throughout the centre but less intense at the edge. C. High power magnification of area indicated in Figure 1F. NCAM intensity was lower at the edges of the blastema (white arrows). D. Quantification of signal intensities for NCAM across the white line in C (blastema) reflects that there was a higher expression of NCAM in the centre of the blastema tissue than at the edges. Scale bars are 50 µm.

### SIX2 and CITED1 co-expression

We analysed co-expression patterns between SIX2 and CITED1, which when co-expressed, are regarded as CSC-associated markers (Petrosyan et al., 2023). CITED1 was expressed in the condensed metanephric mesenchyme and early nephrogenic structures in hFK sections, similar to the SIX2 expression domains (Supplementary Figure 1E, F) (Lovvorn et al., 2007; Murphy et al., 2012). Overall, CITED1 expression was detected in 15 of 18 WT cases (Table 1, Supplementary Table 3, Supplementary Figure 5). By using immunofluorescence in WT specimens, we observed CITED1 expression in matching domains with SIX2 in blastemal component in 14 patients (Figure 1G-J, Table 1). In all samples with CITED1 expression, the protein was localised in the cytoplasm whilst SIX2 protein was always nuclear as previously reported (Murphy et al., 2012). In 5 selected blastemal regions of interest in WT index case 1, we used Pearson’s correlation analysis to display high correlation with a mean of 0.71 (+/- 0.05 SD) of SIX2 and CITED1 co-expression.

### NCAM, SIX2 and PAX2 co-expression

Because NCAM is a marker for multipotent progenitor cells of nephrons and has been reported to identify CSCs (Pode-Shakked et al., 2016; Pode-Shakked et al., 2013), we assessed its expression in WT blastema. In hFK sections, NCAM was expressed in the cap mesenchyme and nephrogenic structures in the renal cortex, as well as in interstitial cells of the kidney medulla (Supplementary Figure 3A). Overall, we detected NCAM expression which was restricted to the blastemal tissue domains, in 15 of 18 WT index cases (Figure 1E, F, Table 1, Supplementary Table 3). In most cases this expression overlapped with SIX2 and PAX2 expression domains (see Figures 1A-F, 2A, C, 3A). These findings are consistent with a previously published study where SIX2 and NCAM were found to be co-expressed in human fetal kidney and naïve WT specimens (Pierce et al., 2014).

**Figure 3.**
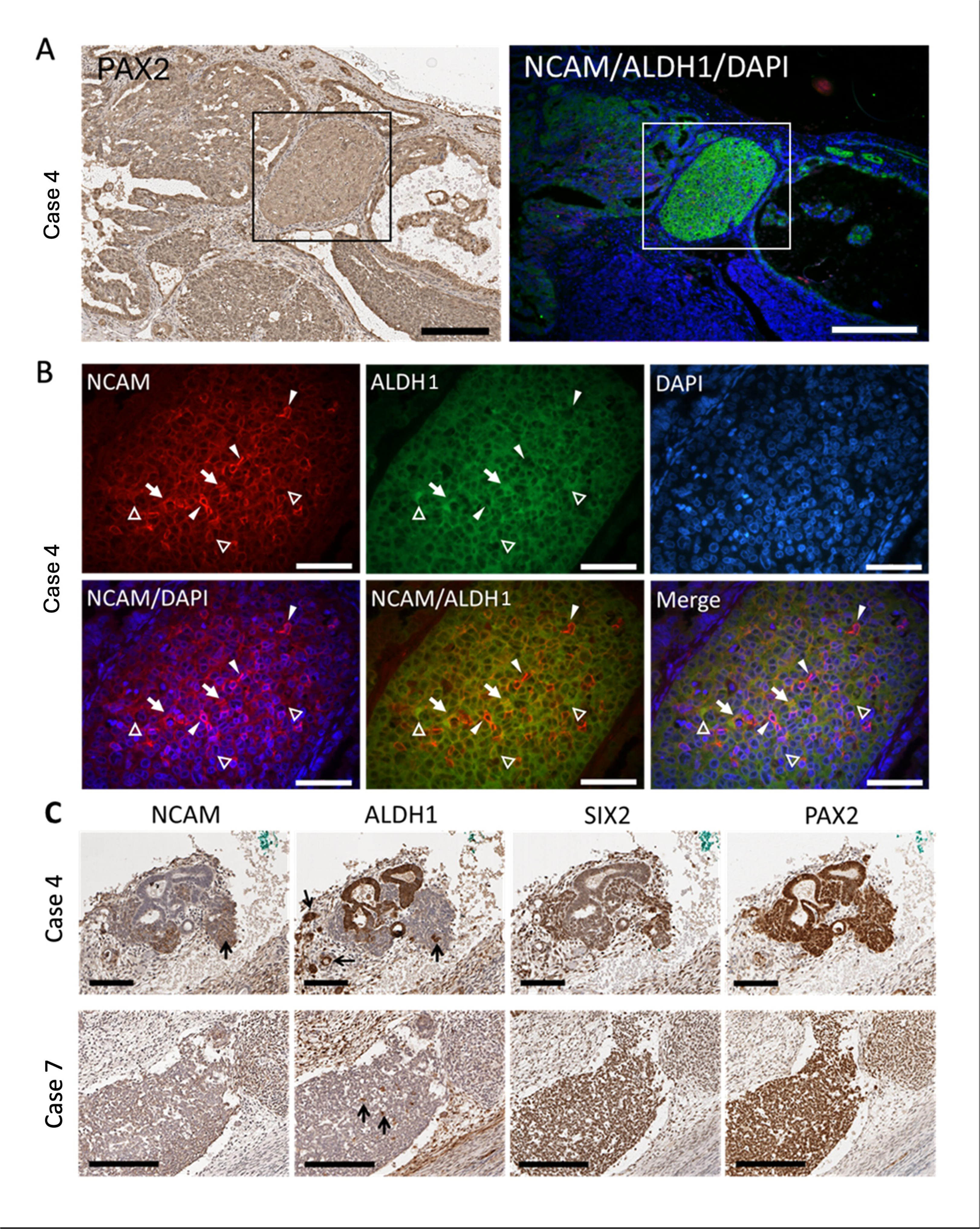
IHC and IF analysis of PAX2, SIX2, NCAM and ALDH1 expression in WT tissue. **A.** In WT index case 4, IHC staining of PAX2 highlights its expression in the blastema region (black square, left), while double IF staining for NCAM (red) and ALDH1 (green) shows their co-localization in the same region (right). DAPI demarcates nuclei in blue. Boxed region shown in B. **B.** Enlarged view of NCAM and ALDH1 co-staining, with the merged image illustrating their spatial relationships. DAPI demarcates nuclei in blue. Arrows point to bright NCAM- and ALDH1-expressing cells, while arrowheads point to bright NCAM-only expressing, and open arrowheads to bright ALDH1-only expressing cells, above the general NCAM- and ALDH1-positivity in the blastema. **C.** IHC staining shows NCAM and ALDH1 expression (black arrows) along with PAX2 and SIX2 expression in epithelial and blastemal tissues in another region in WT index case 4 (upper) and in blastemal tissue in WT index case 7 (lower). Scale bars: 200 µm (A, B, C lower); 90 µm (C upper).

### NCAM and ALDH1 co-expression

Because co-expression of NCAM and ALDH1 have been reported as unequivocally defining WT CSCs (Pode-Shakked et al., 2013; Raved et al., 2019), we further explored their expression patterns. In hFK sections, ALDH1 was detected in mature tubule (epithelial) structures in the renal medulla, in a mutually exclusive pattern to NCAM (Supplementary Figure 3) (Metsuyanim et al., 2009; Pode-Shakked et al., 2009; Raved et al., 2019). Overall, ALDH1 expression was detected in 17 of 18 WT cases (Supplementary Table 3). Co-staining for NCAM and ALDH1 was found in 14 WT tissue specimens (Table 1, Supplementary Figure 5). Among the 14 WT tumours with expression of both NCAM and ALDH1, there were only 4 cases (cases number 4, 7, 11, 17) in which we observed individual cells co-expressing NCAM and ALDH1 in the blastema component (Figure 3, Supplementary Figure 4, Table 1). In WT index case 4, we detected expression of both NCAM and ALDH1 within PAX2-positive blastema tissue. Cells with higher expression intensity for either NCAM or ALDH1 were scattered throughout the blastema, some of which co-expressed high levels of both NCAM and ALDH1 (Figure 3A, B). Similar to our observation in Figure 1E, F and Figure 2C, D, we found that NCAM expression was less intense at the periphery of the blastema. This was in contrast to ALDH1 which was strongly expressed throughout (Figure 3B). In a different region of the same tumour specimen, epithelial structures were situated next to the residual blastema, in a similar pattern to cap mesenchyme and nephrogenic comma- and S-shaped structures (Figure 3C, upper). PAX2 was strongly expressed throughout these cells, while SIX2 was more strongly expressed in the mesenchyme than the epithelial cells. NCAM expression was very similar to SIX2, but ALDH1 expression was strongest in the epithelia and some individual small clusters within the blastema and the stroma. Some of these ALDH1-positive cells were also expressing NCAM. In WT index case 7, SIX2 and PAX2 expression was uniform throughout blastemal structures, which also had uniform NCAM expression (Figure 3C, lower). Interestingly, as in index case 4, there were scattered high-expressing ALDH1-positive cells found across the blastema, which had also otherwise uniform ALDH1 expression at a lower level (Figure 3C lower, compare to Figure 3B).

### CD133 expression

We analysed expression of CD133, since recent literature postulated that it serves as a further marker for WT CSCs (Liu et al., 2018; Singh et al., 2023). We found CD133 was expressed in the cortical nephrogenic zone in comma- and S-shaped bodies in hFK sections (Supplementary Figure 1G, H), in a pattern similar to that observed for PAX2, SIX2, CITED1 and NCAM. This indicates that CD133, like the other progenitor markers, plays a key role in nephrogenic events in the normal kidney, suggesting a role in WT. We explored this further by performing immunolabelling for CD133 in specimens of WT case 7, where we identified its expression near epithelial/tubular structures in nephrogenic rest areas (Figure 4A, B). Notably, WT cases 7 and 17 (Figure 4C, D) were the only two shown to exhibit CD133 positivity throughout the analysed tumour tissues (Table 1).

**Figure 4.**
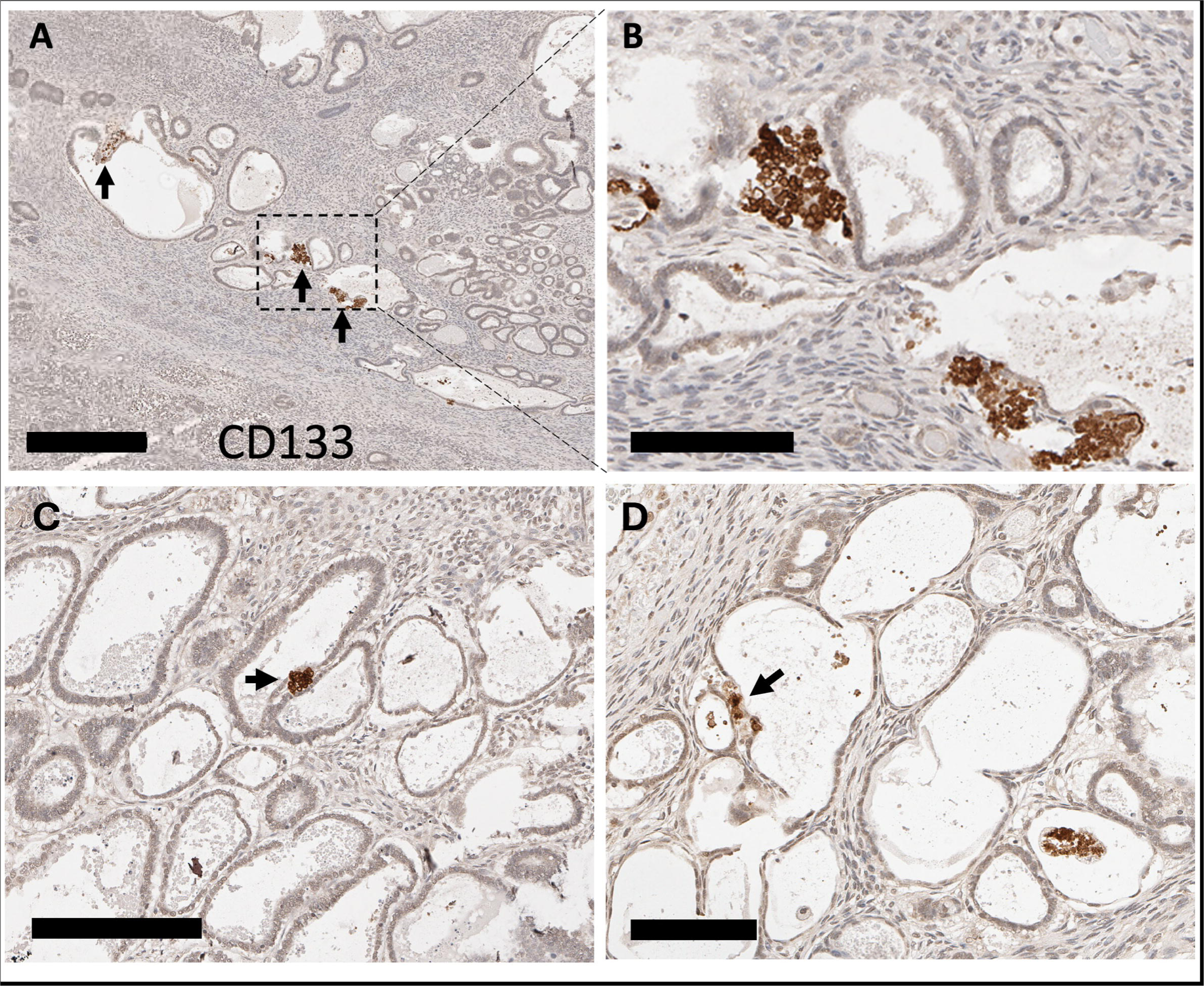
IHC analysis of WT tissue using CD133 marker. **A**, **B.** IHC staining of CD133 of WT tissue in WT index case 7. B shows higher magnification of the boxed area in A. **C, D.** CD133 expression in WT index case 17. Arrows indicate regions with positive staining for CD133. Scale bars are 400 µm (A), 90 µm (B), 200 µm (C) and 100 µm (E).

### Co-expression relationships of kidney progenitor and CSC markers

In order to further evaluate the co-expression relationships between the markers within the 18 WT cases (Table 1), we visualised similarity and expression relationships across blastema, epithelium and stroma using a Jaccard distance-based clustered heat map and Circos plot (Supplementary Figure 5, Figure 5A). The Circos similarity plot revealed strong links in expression between blastemal and epithelial PAX2, blastemal SIX2, CITED1, NCAM and ALDH1 across the WT cases. We also focussed on co-expression relationships solely in the blastema in all 18 WT cases (Supplementary Figure 5), and found the strongest blastemal co-expression between PAX2, SIX2, CITED1 and NCAM (Figure 5B).

**Figure 5.**
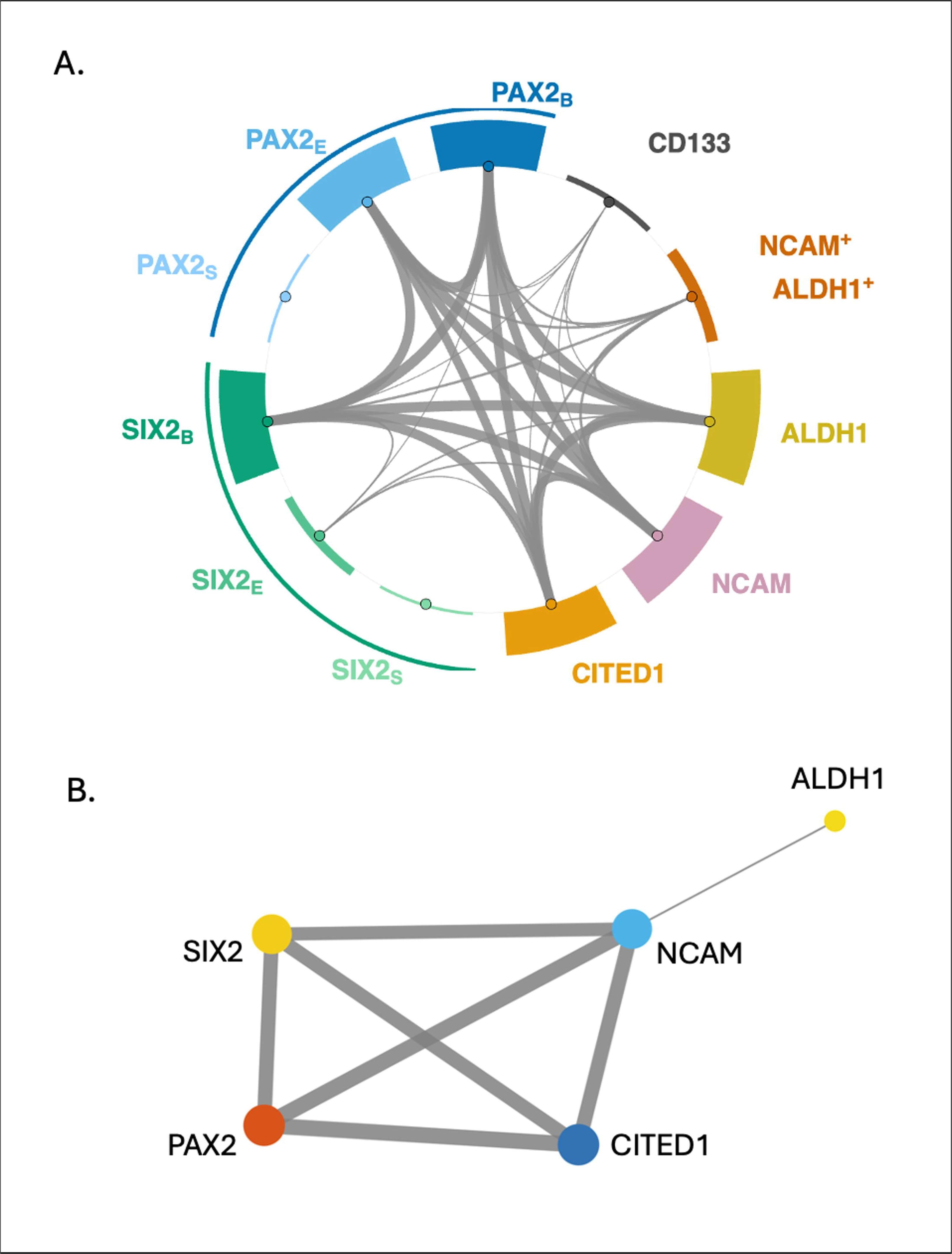
Co-expression relationships between human kidney progenitor markers in WT tissue. A. The Circos-style network illustrates the close spatial relationship between PAX2, SIX2, CITED1, NCAM and ALDH1 expression in the post-chemotherapy WT cases, with the thicker connecting lines illustrating stronger co-expression relationships. B. The network diagram visualises the close co-expression relationship between PAX2, SIX2, CITED1 and NCAM in blastema tissue.

## Discussion

In this study, we have for the first time provided a comprehensive analysis of progenitor and CSC-associated marker expression in 18 WT patients treated with neoadjuvant chemotherapy. We demonstrate the persistence of cells expressing kidney progenitor and CSC-associated markers in the blastema of WT kidneys after standard 4-6 weeks of SIOP-style neoadjuvant chemotherapy. Our results indicate that PAX2, SIX2, CITED1 and NCAM-expressing cells can be detected in the blastema in the majority of cases. Furthermore, we observed co-expression of NCAM and ALDH1 or CD133 in 22% and 11% of cases, respectively; both markers have been previously suggested as strong indicators of CSCs in WT samples from patients treated according to the COG pathway, with upfront nephrectomy followed by neoadjuvant chemotherapy (Jafarian et al., 2020; Mehrazma et al., 2013; Pode-Shakked et al., 2009; Pode-Shakked et al., 2013; Raved et al., 2019; Singh et al., 2023).

One of the key strengths of this current study is the targeted focus on post-chemotherapy WT nephrectomy tissue as per SIOP WT therapy protocols. However, we did not aim to associate our spatial expression analysis with the clinical characteristics or patients outcomes, which we present as a descriptive overview in Supplementary Table 1.

Nevertheless, we would like to point to the fact that two of the 18 patients were deemed to have high risk histological subtypes (blastemal or diffuse anaplasia), and one of these patients (case 7) relapsed. Incidentally, case 7 describes one of 4 patients who had NCAM and ALDH1-co-expression, and also CD133 expression within the tumour. The other patient with high risk histological subtype, case 17, also had NCAM and ALDH1-coexpression and CD133 expression, but is alive without further events. The other patients with co-expression of NCAM and ALDH1 (cases 4 and 11), had stromal or regressive type of histology, suggesting a heterogeneous pattern. However, case numbers are too small to draw any meaningful conclusions.

WT nephrectomy specimens from SIOP-treated patients after having received neo-adjuvant chemotherapy are an underexplored ‘knowledge gap’ when compared to COG studies that entail upfront nephrectomy. While our study is largely observational, it has provided analysis of 18 patients with WT enriched with viable residual blastema providing clear evidence of spatially patterned marker expression, suggesting spatial heterogeneity. A previous study examining marker expression in the blastema of chemotherapy-treated patients following SIOP guidelines confirmed that CITED1 and NCAM as well as the SIX2-related protein SIX1 were expressed in blastemal tissues (Sehic et al., 2014). By contrast, the study by Sehic and colleagues analysed sections from 30 WT patients in a mixed cohort not specifically selected for triphasic characteristics. Furthermore, co-expression analysis was not included in the assessment, nor was there a focus on the localisation of CSC-associated markers.

In our study, hFK samples werved as a critical benchmark for interpreting marker expression. CD133 is a surface marker frequently associated with stem cells in different cancers (Barzegar Behrooz et al., 2019; Glumac and LeBeau, 2018). Analysis of the distribution of CD133 expression in the analysed WT cases showed a highly restricted and unique pattern, being confined to cells near epithelial tumour components and notably absent from the blastema. This finding stands in stark contrast to the high prevalence of CD133 expression reported in the existing literature. For instance, Singh and colleagues (2023) observed CD133 expression in 100% of their cases, while Jafarian and colleagues (2020) and Mehrazma and colleagues (2013) reported positivity rates of in 73.9% and 44%, respectively (Jafarian et al., 2020; Mehrazma et al., 2013; Singh et al., 2023).

The significant discrepancy between our positivity rate of 11% (2/18) and those reported is particularly noteworthy when considering the scale of tissue examined. Unlike studies utilising tissue microarrays or small representative cores, we employed large whole-tissue sections. From a methodological standpoint, larger sections provide a statistically higher probability of identifying rare cell populations. The fact that CD133 remained scarce in our cohort despite this extensive sampling suggests that its low prevalence is a robust biological feature of these samples rather than a result of sampling error.

We propose that this difference is primarily driven by the timing of the tissue collection relative to treatment. While the aforementioned studies often utilized pre-chemotherapy samples or cohorts where treatment status was not specified, our study exclusively analyzed post-chemotherapy tissue. This suggests that the CD133-positive population may be highly sensitive to neoadjuvant chemotherapy and is effectively depleted during treatment. However, it is important to acknowledge a limitation in this interpretation: our cohort did not include paired pre- and post-chemotherapy samples from the same patients. Having access to such paired samples would have allowed for a direct longitudinal assessment of how chemotherapy alters marker expression within the same tumor environment, which would have rendered our interpretation of chemosensitivity even more reliable.

Beyond biological and sampling factors, technical aspects also influence comparative immunohistochemistry. Variability may arise from different antibody clones for CD133, antigen retrieval techniques and tissue processing conditions, particularly duration and quality of formalin fixation, which can significantly affect epitope preservation. Such technical nuances are known confounders in multicentre comparisons and likely contribute to the reported variability.

Ultimately, our findings align with the functional data provided by Pode-Shakked and colleagues (2016), who demonstrated that NCAM+/CD133- cells in WT harbor greater stemness and clonogenic potential than their NCAM+/CD133+ counterparts (Pode-Shakked et al., 2016). When comparing our low post-chemo prevalence of CD133-expressing cells with their findings, CD133+ cells do not appear to be part of the therapy-resistant cancer stem cell population, and to be largely eradicated by standard neoadjuvant regimens. Instead, the persistent, chemotherapy-resistant niches in Wilms tumor appear to be dominated by NCAM and other markers.

Furthermore, in this report, we provide insight into the spatial arrangements of kidney progenitor and CSC-associated marker expression in post-chemotherapy treated WT cases. Our analysis demonstrated recurrent ‘periphery-centre gradients’ within viable blastema tissue where PAX2 expression was high at the periphery, and NCAM expression was highest within the centre regions studied. The variation in PAX2 intensity, stronger at the periphery, suggests a dynamic expression landscape, where peripheral cells may be transitioning toward epithelial differentiation via mesenchymal-to-epithelial transition (MET), while central blastemal cells retain mesenchymal progenitor features. This spatial arrangement mirrors differentiation hierarchies in normal fetal kidney development in the cap mesenchyme, suggesting that MET is active in chemotherapy-treated WT blastema tissue. A recent study using human kidney organoids showed that MET events are regulated by PAX8 and that the closely related PAX2 is expressed in uncommitted and committed cap mesenchyme and the subsequently committed epithelium (renal vesicle, comma- and S-shaped bodies) (Ng-Blichfeldt et al., 2024). These findings match our own data presented here. NCAM, also known as CD56, is a cell adhesion molecule pivotal in cell-to-cell interactions. NCAM staining in hFK has previously been demonstrated in the cap mesenchyme and uncommitted epithelium, and undifferentiated blastema in untreated WT (Buzhor et al., 2013; Kapur and Rakheja, 2011; Metsuyanim et al., 2009; Pode-Shakked et al., 2016). In WT, NCAM expression has been associated with tumour progression, metastasis, and ‘stemness properties’ (Pode-Shakked et al., 2009; Pode-Shakked et al., 2013; Raved et al., 2019). As discussed above, Pode-Shakked and colleagues concluded that NCAM-expressing blastemal mesenchyme cells within untreated WT tissue are a likely source of CSCs (Pode-Shakked et al., 2016).

In our current study, we provide evidence of the co-expression of PAX2, SIX2, CITED1, ALDH1 and NCAM1 with distinct expression localisation, mostly in blastema. All of these proteins play a role in normal human kidney development. Our results showed that PAX2 and SIX2 were co-expressed in the blastema in 83% of WT cases, supporting their pivotal roles in kidney progenitor-like cells. PAX2 was present in both epithelial and blastemal elements, with occasional stromal localisation, highlighting its broader expression profile compared to SIX2, which was largely confined to blastemal cells, consistent with maintaining progenitor status. SIX2 in combination with WNT signalling and via *β*-Catenin activity, regulates the self-renewal of nephron progenitor cells within the condensed cap mesenchyme of the developing nephrons in normal kidney development (Kobayashi et al., 2008; Park et al., 2012; Self et al., 2006).

SIX2 and CITED1 showed overlapping expression in WT blastema. Distinct subcellular localisation (nuclear SIX2 and cytoplasmic CITED1 (Murphy et al., 2012)) suggests complementary functions, with SIX2 regulating transcriptional self-renewal and CITED1 modulating progenitor signalling. CITED1 is an early marker for undifferentiated metanephric mesenchyme which gives rise to nephron structures and is co-expressed with SIX2 in nephron progenitor cells during normal development and in WT (Boyle et al., 2008; Lindstrom et al., 2018; Murphy et al., 2012). In an in vitro system, CITED1 was found to convey ‘stemness’ to WT cells when forced to localise to the nucleus (Murphy et al., 2014). Finally, a recent study identified SIX2-, CITED1-coexpressing cells as CSCs in WT using spatial transcriptomics (Petrosyan et al., 2023). The persistence of SIX2-,CITED1-positive cells in post-therapy WT suggests that progenitor-like states are retained despite treatment. In untreated WT, these double-positive cells have been validated as CSCs with tumour-propagating potential (Petrosyan et al., 2023), suggesting they may represent a therapy-resistant, CSC-enriched population contributing to disease relapse.

The significance of cells expressing high levels of NCAM and ALDH1 remains unclear. Previous studies reported that ALDH1-expressing cells within NCAM positive blastema defined WT CSCs (Pode-Shakked et al., 2013; Raved et al., 2019), and xenotransplantation confirmed ALDH1 as marker for tumour-propagating cells (Shukrun et al., 2014). NCAM/ALDH1-positive blastemal cells correlate with poor outcomes, whereas epithelial or stromal double-positive cells do not (Raved et al., 2019). ALDH1 is an enzyme that participates in the oxidation of aldehydes and is involved in various cellular processes, including detoxification. Therefore, ALDH1 in combination with NCAM expression in blastema tissue suggests that it plays a role in protecting cancer cells against cytotoxic agents including chemotherapy in WT (Pode-Shakked et al., 2013). Co-expression of both markers in WT cells was shown to be associated with significant elevation in the transcript expression of early renal progenitor markers *SIX2*, *PAX2*, *SALL1*, and *OSR1*, indicating that these cells represent CSCs (Pode-Shakked et al., 2016; Pode-Shakked et al., 2013). Further experimental and clinical analyses have revealed an association of NCAM- and ALDH1-co-expressing cells with a more aggressive tumour phenotype and poor prognosis, enhanced tumour-initiating potential, and resistance to chemotherapy (Raved et al., 2019). Therefore, our finding of NCAM and ALDH1 co-expressing cells in the blastemal component of post-chemotherapy WT supports the key observations that the blastema retains embryonic kidney progenitor- and/or WT cancer stem cells (Pop et al., 2025).

CSCs in WT have also been characterised by co-expression of NCAM and SIX2 (Pierce et al., 2014; Pode-Shakked et al., 2013; Raved et al., 2019). Pierce and colleagues showed that SIX2-and NCAM-co-expression was mostly found in WT blastema tissue and also observed co-expression of NCAM with CITED1 (Pierce et al., 2014).

It is important to note that the functional significance of all these markers may vary between individuals and different stages of WT (Spreafico et al., 2021).

## Limitations

We acknowledge that the small number of post-chemotherapy WT cases examined in this study does not allow us to establish a statistical correlation between marker expression and WT staging or disease relapse prediction, and thus limits our analyses as exploratory and descriptive. A larger WT cohort will be needed to further test these relationships and their underlying causation.

Our results describe the marker expression landscape with respect to kidney progenitor and CSC-associated markers, with a particular focus on WT with viable blastema. Our data may therefore be affected by selection bias since other WT subtypes such as diffuse anaplasia and WT with WT1 mutations may yet present different marker landscapes. Furthermore, entrapped nephrogenic rests could be interpreted as having progenitor signatures.

It is important to also note that chemotherapy may potentially induce reprogramming of tumour cells, selecting for NCAM and ALDH1 states and hence change the existing progenitor and stem cell signatures within neoplatstic tissue (Ayub et al., 2015; Blaheta et al., 2006).

The descriptive nature of this study precludes us from unequivocally stating that cells with CSC-associated expression signatures are indeed CSCs. Further functional analyses will be needed to fully determine the nature of these cells, including xenograft studies and functional assays.

## Conclusion

Taken together, our findings have provided insight into the co-expression and spatial arrangement of kidney progenitor and CSC markers within WT specimens containing viable residual blastema tissue, following neo-adjuvant chemotherapy. Our data suggest that the pre-treated WT specimens have similar expression patterns of progenitor/CSC markers within the blastema when compared to naïve WT specimens. Our study provides evidence that there is a strong expression overlap between PAX2, SIX2, CITED1, NCAM and ALDH1 within pre-treated blastema, and that SIX2, CITED1 and NCAM are co-expressed, indicating that cells with nephron progenitor characteristics persist in complex patterns. Since co-expression of SIX2, CITED1, NCAM and ALDH1 have been suggested to indicate WT CSCs, our data suggest that CSC-associated cells may be present in the blastema of pre-treated WT and in close vicinity to nephron progenitor cells. Further analysis is required to establish a deeper understanding of spatially organised progenitor-like niches in WT that persist following chemotherapy, including the nuances of cell-cell interactions.

## Data Availability

All research data are available from the corresponding author on reasonable request.

## Acknowledgements

WT nephrectomy specimens and datasets used in this study were kindly provided by UK VIVO Biobank, supported by Cancer Research UK & Blood Cancer UK (Grant no. CRCPSC-Dec21\100003), and the UK IMPORT study currently funded by the Children’s Cancer and Leukaemia Group (CCLG)/Little Princess Trust (Grant ref: CCLGA 2019 27) and previously funded by CCLG/Bethany’s Wish (grant ref: CCLGA 2017 02), EU-FP7 grant refs: 261474 (ENCCA) and 270089 (P-medicine), Great Ormond Street Children’s Charity (grant ref: W1090) and Cancer Research UK (grant ref: C1188/A17297). The study received infrastructural support from the UK National Cancer Research Network and the CCLG. We thank Dr Kathy Pritchard-Jones, Dr Reem Al-Saadi, Dr Tanzina Chowdhury as past and present CIs of the IMPORT study for their support of this work.

We thank the CCLG VIVO Biobank, UK CCLG centres, the UK IMPORT study and the ECMC Paediatric Network for consenting the patients and the collection and provision of the WT tissue samples. We extend gratitude and thanks to all patients and families for collaboration and tissue specimen donation for the research.

We acknowledge expert technical assistance provided by the University of Liverpool Biobank for generating tissue sections, and for scanning stained specimen slides, and the University of Liverpool LIV-SRF Histology Facility for expert support in generating tissue sections and cover slipping slides.

## Ethics statement

Parent/guardian informed consent was in place for the specimens collected from 18 WT index patients and the human foetal kidney specimen. Ethical approval of the study was by the UK VIVO Biobank (project number 23-VIVO-10) and the UK IMPORT study for renal tumours of childhood (REC reference 12/LO/0101), and for the foetal human kidney specimen under REC reference 25/PR/0493. Institutional sponsorship was granted for this study. The study followed all applicable regulations and ethical standards for the use of human tissue in biomedical research and adhered to the principles of the Declaration of Helsinki, and observed the privacy rights of human subjects.

## Author Contribution

MM designed and performed the experiments, analysed the data, curated the data, prepared the figures and wrote the draft and final version of the manuscript. LH, BP, PL obtained funding, provided clinical input as medical oncologists and surgeons, and edited and contributed to the final manuscript version. PM obtained funding, provided advice and contributed to the final manuscript version. SA provided clinical data as pathologist, edited and contributed to the final manuscript version. EC and RS obtained funding, provided samples, clinical advice as pathologists, and edited and contributed to the final manuscript version. BW obtained funding, designed experiments, supervised and managed the project, prepared the figures, wrote the draft and final version of the manuscript.

## Declaration of generative AI and AI-assisted technologies in the manuscript preparation process

During the preparation of this manuscript, the authors did not use any AI tools.

## Availability of data

All research data are available from the corresponding author on reasonable request.

## References

Ayub, T. H., Keyver-Paik, M. D., Debald, M., Rostamzadeh, B., Thiesler, T., Schroder, L., Barchet, W., Abramian, A., Kaiser, C., Kristiansen, G., et al. (2015). Accumulation of ALDH1-positive cells after neoadjuvant chemotherapy predicts treatment resistance and prognosticates poor outcome in ovarian cancer. Oncotarget 6, 16437–16448.

Barzegar Behrooz, A., Syahir, A. and Ahmad, S. (2019). CD133: beyond a cancer stem cell biomarker. J Drug Target 27, 257–269.

Blaheta, R. A., Daher, F. H., Michaelis, M., Hasenberg, C., Weich, E. M., Jonas, D., Kotchetkov, R., Doerr, H. W. and Cinatl, J., Jr. (2006). Chemoresistance induces enhanced adhesion and transendothelial penetration of neuroblastoma cells by down-regulating NCAM surface expression. BMC Cancer 6, 294.

Boyle, S., Misfeldt, A., Chandler, K. J., Deal, K. K., Southard-Smith, E. M., Mortlock, D. P., Baldwin, H. S. and de Caestecker, M. (2008). Fate mapping using Cited1-CreERT2 mice demonstrates that the cap mesenchyme contains self-renewing progenitor cells and gives rise exclusively to nephronic epithelia. Dev Biol 313, 234–245.

Brok, J., Lopez-Yurda, M., Tinteren, H. V., Treger, T. D., Furtwängler, R., Graf, N., Bergeron, C., van den Heuvel-Eibrink, M. M., Pritchard-Jones, K., Olsen, Ø. E., et al. (2018). Relapse of Wilms’ tumour and detection methods: a retrospective analysis of the 2001 Renal Tumour Study Group–International Society of Paediatric Oncology Wilms’ tumour protocol database. The Lancet Oncology 19, 1072–1081.

Buzhor, E., Omer, D., Harari-Steinberg, O., Dotan, Z., Vax, E., Pri-Chen, S., Metsuyanim, S., Pleniceanu, O., Goldstein, R. S. and Dekel, B. (2013). Reactivation of NCAM1 defines a subpopulation of human adult kidney epithelial cells with clonogenic and stem/progenitor properties. The American journal of pathology 183, 1621–1633.

Dickinson, K., Bernard, C., Iglesias, D. and Goodyer, P. (2022). Nephron progenitor cells in development and renal disease: Renal Hypoplasia, Wilms Tumour and recovery from Acute Kidney Injury. Medical Research Archives; Vol 10 No 1 (2022): Vol.10 Issue 1, January, 2022.

Fawkner-Corbett, D. W., Howell, L., Pizer, B. L., Dominici, C., McDowell, H. P. and Losty, P. D. (2014). Wilms’ tumor--lessons and outcomes--a 25-year single center UK experience. Pediatr Hematol Oncol 31, 400–408.

Glumac, P. M. and LeBeau, A. M. (2018). The role of CD133 in cancer: a concise review. Clinical and translational medicine 7, 18.

Groenendijk, A., Spreafico, F., de Krijger, R. R., Drost, J., Brok, J., Perotti, D., van Tinteren, H., Venkatramani, R., Godzinski, J., Rube, C., et al. (2021). Prognostic Factors for Wilms Tumor Recurrence: A Review of the Literature. Cancers (Basel) 13.

Jafarian, A. H., Zabolinejad, N., Mohamadian Roshan, N., Hashemi, S. and Gharib, M. (2020). Evaluation of CD133 and CD56/NCAM expression in Wilms tumor and their association with prognostic factors. Iranian journal of basic medical sciences 23, 853–857.

Kapur, P. and Rakheja, D. (2011). Immunohistochemical expression of neural cell adhesion molecule in Wilms tumors, nephrogenic rests, and fetal and postnatal renal cortices. Pediatr Dev Pathol 14, 16–19.

Khoshdel Rad, N., Aghdami, N. and Moghadasali, R. (2020). Cellular and Molecular Mechanisms of Kidney Development: From the Embryo to the Kidney Organoid. Frontiers in Cell and Developmental Biology 8.

Kobayashi, A., Valerius, M. T., Mugford, J. W., Carroll, T. J., Self, M., Oliver, G. and McMahon, A. P. (2008). Six2 defines and regulates a multipotent self-renewing nephron progenitor population throughout mammalian kidney development. Cell stem cell 3, 169–181.

Li, H., Hohenstein, P. and Kuure, S. (2021). Embryonic Kidney Development, Stem Cells and the Origin of Wilms Tumor. Genes 12, 318.

Lindstrom, N. O., Guo, J., Kim, A. D., Tran, T., Guo, Q., De Sena Brandine, G., Ransick, A., Parvez, R. K., Thornton, M. E., Baskin, L., et al. (2018). Conserved and Divergent Features of Mesenchymal Progenitor Cell Types within the Cortical Nephrogenic Niche of the Human and Mouse Kidney. Journal of the American Society of Nephrology : JASN 29, 806–824.

Losty, P. D. (2016). Evidence-based paediatric surgical oncology. Seminars in Pediatric Surgery 25, 333–335.

Lovvorn, H. N., Westrup, J., Opperman, S., Boyle, S., Shi, G., Anderson, J., Perlman, E. J., Perantoni, A. O., Wills, M. and de Caestecker, M. (2007). CITED1 expression in Wilms’ tumor and embryonic kidney. Neoplasia 9, 589–600.

Marzagalli, M., Fontana, F., Raimondi, M. and Limonta, P. (2021). Cancer Stem Cells-Key Players in Tumor Relapse. Cancers (Basel) 13.

Mehrazma, M., Madjd, Z., Kalantari, E., Panahi, M., Hendi, A. and Shariftabrizi, A. (2013). Expression of stem cell markers, CD133 and CD44, in pediatric solid tumors: a study using tissue microarray. Fetal Pediatr Pathol 32, 192–204.

Metsuyanim, S., Harari-Steinberg, O., Buzhor, E., Omer, D., Pode-Shakked, N., Ben-Hur, H., Halperin, R., Schneider, D. and Dekel, B. (2009). Expression of stem cell markers in the human fetal kidney. PLoS One 4, e6709.

Murphy, A. J., Pierce, J., de Caestecker, C., Ayers, G. D., Zhao, A., Krebs, J. R., Saito-Diaz, V. K., Lee, E., Perantoni, A. O., de Caestecker, M. P., et al. (2014). CITED1 confers stemness to Wilms tumor and enhances tumorigenic responses when enriched in the nucleus. Oncotarget 5, 386–402.

Murphy, A. J., Pierce, J., de Caestecker, C., Taylor, C., Anderson, J. R., Perantoni, A. O., de Caestecker, M. P. and Lovvorn, H. N., 3rd (2012). SIX2 and CITED1, markers of nephronic progenitor self-renewal, remain active in primitive elements of Wilms’ tumor. J Pediatr Surg 47, 1239–1249.

Ng-Blichfeldt, J. P., Stewart, B. J., Clatworthy, M. R., Williams, J. M. and Roper, K. (2024). Identification of a core transcriptional program driving the human renal mesenchymal-to-epithelial transition. Dev Cell 59, 595–612 e598.

Ortiz, M. V., Koenig, C., Armstrong, A. E., Brok, J., de Camargo, B., Mavinkurve-Groothuis, A. M. C., Herrera, T. B. V., Venkatramani, R., Woods, A. D., Dome, J. S., et al. (2023). Advances in the clinical management of high-risk Wilms tumors. Pediatr Blood Cancer 70 Suppl 2, e30342.

Park, J. S., Ma, W., O’Brien, L. L., Chung, E., Guo, J. J., Cheng, J. G., Valerius, M. T., McMahon, J. A., Wong, W. H. and McMahon, A. P. (2012). Six2 and Wnt regulate self-renewal and commitment of nephron progenitors through shared gene regulatory networks. Dev Cell 23, 637–651.

Pasqualini, C., Furtwangler, R., van Tinteren, H., Teixeira, R. A. P., Acha, T., Howell, L., Vujanic, G., Godzinski, J., Melchior, P., Smets, A. M., et al. (2020). Outcome of patients with stage IV high-risk Wilms tumour treated according to the SIOP2001 protocol: A report of the SIOP Renal Tumour Study Group. Eur J Cancer 128, 38–46.

Petrosyan, A., Villani, V., Aguiari, P., Thornton, M. E., Wang, Y., Rajewski, A., Zhou, S., Cravedi, P., Grubbs, B. H., De Filippo, R. E., et al. (2023). Identification and Characterization of the Wilms Tumor Cancer Stem Cell. Adv Sci (Weinh) 10, e2206787.

Philchenkov, A. and Dubrovska, A. (2024). Cancer Stem Cells as a Therapeutic Target: Current Clinical Development and Future Prospective. Stem cells 42, 173–199.

Pierce, J., Murphy, A. J., Panzer, A., de Caestecker, C., Ayers, G. D., Neblett, D., Saito-Diaz, K., de Caestecker, M. and Lovvorn, H. N., 3rd (2014). SIX2 Effects on Wilms Tumor Biology. Transl Oncol 7, 800–811.

Pode-Shakked, N., Metsuyanim, S., Rom-Gross, E., Mor, Y., Fridman, E., Goldstein, I., Amariglio, N., Rechavi, G., Keshet, G. and Dekel, B. (2009). Developmental tumourigenesis: NCAM as a putative marker for the malignant renal stem/progenitor cell population. J Cell Mol Med 13, 1792–1808.

Pode-Shakked, N., Pleniceanu, O., Gershon, R., Shukrun, R., Kanter, I., Bucris, E., Pode-Shakked, B., Tam, G., Tam, H., Caspi, R., et al. (2016). Dissecting Stages of Human Kidney Development and Tumorigenesis with Surface Markers Affords Simple Prospective Purification of Nephron Stem Cells. Sci Rep 6, 23562.

Pode-Shakked, N., Shukrun, R., Mark-Danieli, M., Tsvetkov, P., Bahar, S., Pri-Chen, S., Goldstein, R. S., Rom-Gross, E., Mor, Y., Fridman, E., et al. (2013). The isolation and characterization of renal cancer initiating cells from human Wilms’ tumour xenografts unveils new therapeutic targets. EMBO Mol Med 5, 18–37.

Pop, N. S., Koot, D., Brouwers, C. M., Linssen, M. M., Claassens, J. W. C., Cartlidge, C. W. J., Özdemir, D. D., Dolt, K. S. and Hohenstein, P. (2025). Genotype-phenotype correlations and de novo induction of cancer stem cells in Wilms tumor initiation. bioRxiv doi: 10.1101/2025.11.19.689177

Popov, S. D., Sebire, N. J. and Vujanic, G. M. (2016). Wilms’ Tumour - Histology and Differential Diagnosis. In Wilms Tumor (ed. M. M. van den Heuvel-Eibrink). Brisbane (AU).

Raved, D., Tokatly-Latzer, I., Anafi, L., Harari-Steinberg, O., Barshack, I., Dekel, B. and Pode-Shakked, N. (2019). Blastemal NCAM(+)ALDH1(+) Wilms’ tumor cancer stem cells correlate with disease progression and poor clinical outcome: A pilot study. Pathol Res Pract 215, 152491.

Royer-Pokora, B., Wruck, W., Adjaye, J. and Beier, M. (2023). Gene expression studies of WT1 mutant Wilms tumor cell lines in the frame work of published kidney development data reveals their early kidney stem cell origin. PLoS One 18, e0270380.

Sehic, D., Ciornei, C. D. and Gisselsson, D. (2014). Evaluation of CITED1, SIX1, and CD56 protein expression for identification of blastemal elements in Wilms tumor. Am J Clin Pathol 141, 828–833.

Self, M., Lagutin, O. V., Bowling, B., Hendrix, J., Cai, Y., Dressler, G. R. and Oliver, G. (2006). Six2 is required for suppression of nephrogenesis and progenitor renewal in the developing kidney. EMBO J 25, 5214–5228.

Shukrun, R., Pode-Shakked, N., Pleniceanu, O., Omer, D., Vax, E., Peer, E., Pri-Chen, S., Jacob, J., Hu, Q., Harari-Steinberg, O., et al. (2014). Wilms’ tumor blastemal stem cells dedifferentiate to propagate the tumor bulk. Stem Cell Reports 3, 24–33.

Singh, S., Bhardwaj, M., Sen, A., Nambiyar, K. and Ahuja, A. (2023). Cancer Stem Cell Markers - CD133 and CD44 - in Paediatric Solid Tumours: A Study of Immunophenotypic Expression and Correlation with Clinicopathological Parameters. Indian J Surg Oncol 14, 113–121.

Spreafico, F., Fernandez, C. V., Brok, J., Nakata, K., Vujanic, G., Geller, J. I., Gessler, M., Maschietto, M., Behjati, S., Polanco, A., et al. (2021). Wilms tumour. Nat Rev Dis Primers 7, 75.

Spreafico, F., Pritchard Jones, K., Malogolowkin, M. H., Bergeron, C., Hale, J., de Kraker, J., Dallorso, S., Acha, T., de Camargo, B., Dome, J. S., et al. (2009). Treatment of relapsed Wilms tumors: lessons learned. Expert Review of Anticancer Therapy 9, 1807–1815.

van den Heuvel-Eibrink, M. M., Hol, J. A., Pritchard-Jones, K., van Tinteren, H., Furtwängler, R., Verschuur, A. C., Vujanic, G. M., Leuschner, I., Brok, J., Rübe, C., et al. (2017). Rationale for the treatment of Wilms tumour in the UMBRELLA SIOP–RTSG 2016 protocol. Nature Reviews Urology 14, 743–752.

van den Heuvel-Eibrink, M. M., van Tinteren, H., Bergeron, C., Coulomb-L’Hermine, A., de Camargo, B., Leuschner, I., Sandstedt, B., Acha, T., Godzinski, J., Oldenburger, F., et al. (2015). Outcome of localised blastemal-type Wilms tumour patients treated according to intensified treatment in the SIOP WT 2001 protocol, a report of the SIOP Renal Tumour Study Group (SIOP-RTSG). Eur J Cancer 51, 498–506.

Vujanic, G. M., Parsons, L. N., D’Hooghe, E., Treece, A. L., Collini, P. and Perlman, E. J. (2022). Pathology of Wilms’ tumour in International Society of Paediatric Oncology (SIOP) and Children’s oncology group (COG) renal tumour studies: Similarities and differences. Histopathology 80, 1026–1037.

