## Supplementary data for "Cancer Stem Cell–Associated Marker Expression in Chemotherapy-Treated Wilms Tumour"

**Supplementary Table 1: Primary Wilms Tumour samples and patient characteristics.**

| Case | Stage at diagnosis | Histological diagnosis | Local stage | Treatment | Events / event free survival |
|---|---|---|---|---|---|
| 1 | localised | Regressive Type | 3 | Pre-op AV / Post-op AV2 + radiotherapy | EF - Alive |
| 2 | localised | Regressive Type | 2 | Pre-op AV / Post-op AV2 | EF - Alive |
| 3 | Stage 4 | Regressive Type | 3 | Pre-op AVD / Post-op HR + radiotherapy | EF - Alive |
| 4 | Stage 4 | Regressive Type | 1 | Pre-op AVD / Post-op AVD | EF - Alive |
| 5 | Stage 4 | Mixed Type | 2 | Pre-op AVD / Post-op HR | Relapse - Alive |
| 6 | Stage 5 | Mixed Type | 1 | Pre-op AV / Post-op AV1 | EF - Alive |
| 7 | Stage 4 | Diffuse anaplasia | 1 | Pre-op AV / Post-op HR | Relapse - Alive |
| 8 | localised | Mixed Type | 2 | Pre-op AV / Post-op AV2 | EF - Alive |
| 9 | localised | Mixed Type | 3 | Pre-op AV / Post-op AV2 + radiotherapy | EF - Alive |
| 10 | Stage 4 | Mixed Type | 2 | Pre-op AV / Post-op HR | EF - Alive |
| 11 | localised | Stromal | 2 | Pre-op AV / Post-op AV2 | EF - Alive |
| 12 | localised | Regressive | 1 | Pre-op AV / Post-op AV1 | EF - Alive |
| 13 | Stage 4 | Epithelial | 3 | Pre-op AVD / Post-op HR + radiotherapy | EF - Alive |
| 14 | localised | Stromal | 1 | Pre-op AV / Post-op AV1 | EF - Alive |
| 15 | localised | Regressive Type | 1 | Pre-op AV / Post-op AV1 | EF - Alive |
| 16 | localised | Regressive Type | 1 | Pre-op AV / Post-op AV2 | EF - Alive |
| 17 | localised | Blastemal Type | 3 | Pre-op AV / Post-op HR + radiotherapy | EF - Alive |
| 18 | localised | Mixed Type | 2 | Pre-op AV / Post-op AV + radiotherapy | EF - Alive |

**AV (Pre-op) / AV1 (Post-op) = 4 weeks Actinomycin D + Vincristine**

**AV2 = 27 weeks Actinomycin D + Vincristine**

**Pre-op AVD = 6 weeks Actinomycin D + Vincristine + Doxorubicin**

**Post-op AVD = 27 weeks Actinomycin D + Vincristine + Doxorubicin**

**Post-op HR = 36 weeks high risk (4 drug) chemotherapy; Cyclophosphamide, Doxorubicin, Carboplatin and Etoposide**

**Pre-op = treatment pre-nephrectomy**

**Post-op = treatment post-nephrectomy**

**EF = Event Free**

**Supplementary Table 2: List of primary and secondary antibodies**

| **Primary antibodies** | | | |
|---|---|---|---|
| **Antibody** | **Source** | **Product code** | **Dilution** |
| PAX2 | Rabbit | 201002 | 1:200 for IF<br>1:1000 for IHC |
| SIX2 | Mouse | 66347-1-Ig | 1:200 for IF<br>1:200 for IHC |
| CITED1 | Rabbit | 26999-1-AP | 1:200 for IF<br>1:1000 for IHC |
| NCAM1 | Mouse | Sc-106 | 1:100 for IF<br>1:200 for IHC |
| ALDH1 | Rabbit | 15910-1-AP | 1:100 for IF<br>1:1000 for IHC |
| CD133 | Rabbit | 18470-1-AP | 1:200 for IHC |
| Secondary antibodies | | | |
| **Antibody** | **Detection** | **Product code** | **Dilution** |
| Goat anti-Mouse IgG2b | Alexa594 | A-21145<br>Invitrogen | 1:1000 |
| Chicken anti-Rabbit IgG | Alexa488 | A-21441<br>Invitrogen | 1:1000 |
| Horse anti-Rabbit IgG | Horseradish Peroxidase | MP-7401<br>Vector laboratories | ImmPRESS kit |
| Horse anti-Mouse IgG | Horseradish Peroxidase | MP-7402<br>Vector laboratories | ImmPRESS kit |

**Supplementary Table 3. H-scores of analysed markers PAX2, SIX2, CITED1, NCAM and ALDH1 in sections of all 18 cases, and summary of incidence.**

| Case | PAX2 | SIX2 | CITED1 | NCAM | ALDH1 |
|---|---|---|---|---|---|
| 1 | 245 | 210 | 275 | 45 | 220 |
| 2 | 260 | 55 | 230 | 90 | 210 |
| 3 | 280 | 180 | 250 | 205 | 150 |
| 4 | 290 | 240 | 280 | 220 | 230 |
| 5 | 210 | 215 | 150 | 250 | 220 |
| 6 | 275 | 265 | 290 | 230 | 210 |
| 7 | 285 | 230 | 260 | 245 | 220 |
| 8 | 220 | 235 | 255 | 210 | 85 |
| 9 | 70 | 150 | 260 | 220 | 240 |
| 10 | 215 | 225 | 90 | 230 | 220 |
| 11 | 275 | 245 | 270 | 240 | 230 |
| 12 | 265 | 220 | 275 | 250 | 210 |
| 13 | 280 | 215 | 260 | 160 | 225 |
| 14 | 150 | 220 | 250 | 240 | 230 |
| 15 | 260 | 240 | 270 | 225 | 160 |
| 16 | 245 | 225 | 180 | 235 | 220 |
| 17 | 270 | 230 | 160 | 240 | 210 |
| 18 | 230 | 90 | 80 | 60 | 210 |

| N = 18 cases | PAX2 | SIX2 | CITED1 | NCAM | ALDH1 |
|---|---|---|---|---|---|
| Positive (H-Score ≥ 101) (percentage of cases) | 17 / 18 (94.4%) | 16 / 18 (88.9%) | 16 / 18 (88.9%) | 15 / 18 (83.3%) | 17 / 18 (94.4%) |
| Positive (H-Score 0-100) (percentage of cases) | 1 / 18 (5.6%) | 2 / 18 (11.1%) | 2 / 18 (11.1%) | 3 / 18 (16.7%) | 1 / 18 (5.6%) |

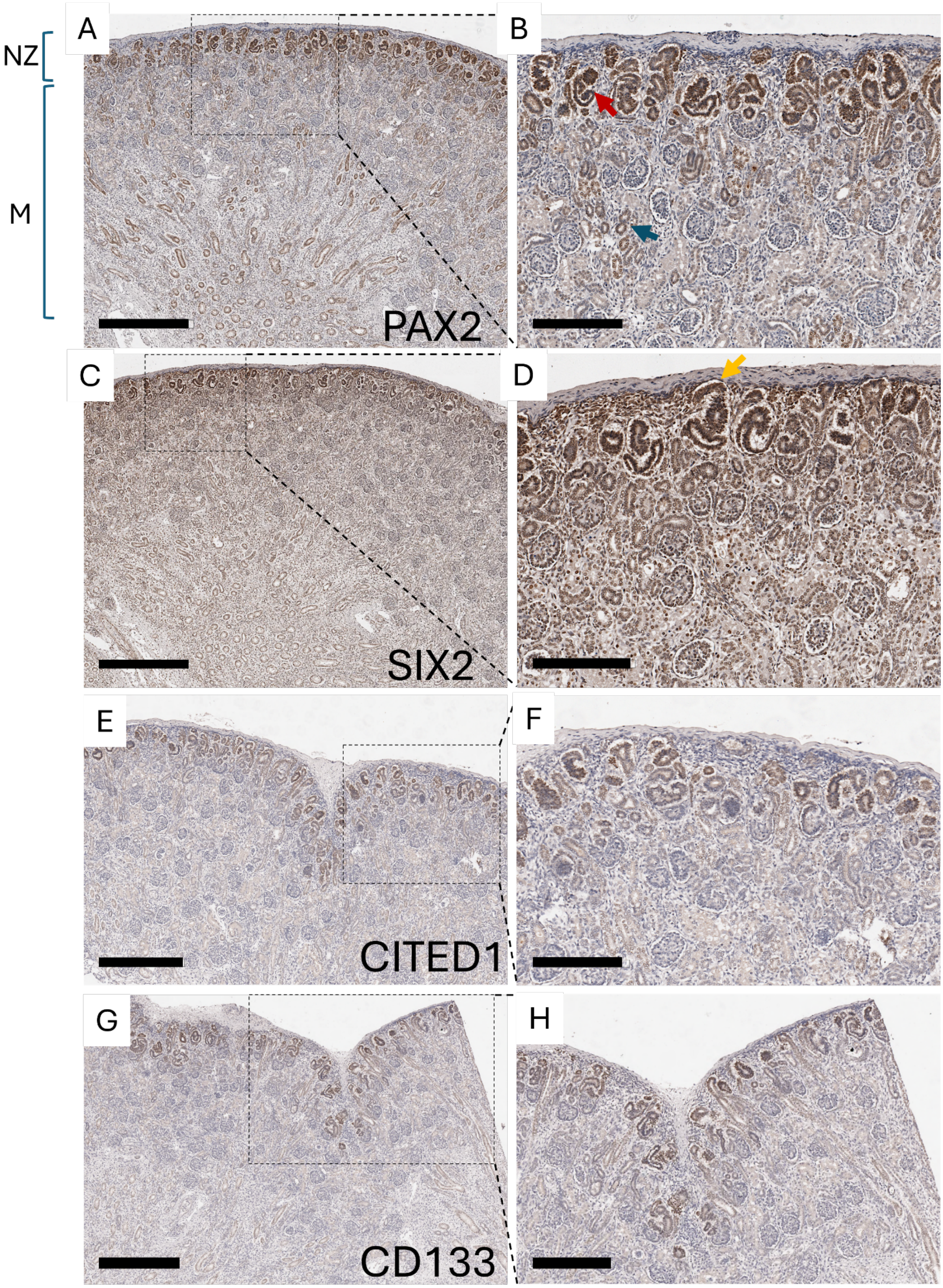


**Supplementary Figure 1. Expression domains of PAX2, SIX2, CITED1 and CD133 in hFK**. Representative IHC of PAX2, SIX2, CITED1 and CD133 on paraffin embedded sections of hFK. **A,**

**B**. PAX2 expression was found within the nephrogenic zone (NZ), containing progenitor structures such as comma-shaped and S-shaped bodies (red arrow). In the medulla, PAX2 expression was limited to immature tubules (blue arrow). **C, D.** SIX2 expression was observed in the cap mesenchyme (orange arrow) and early post- MET structures, including comma-shaped and S-shaped bodies. **E, F.** CITED1 expression was confined to the nephrogenic zone (NZ). **G, H.** CD133 expression was detected not only in early progenitor structures but also in more mature tubules within the medulla. NZ-Nephrogenic zone; M-Medulla. Scale bars are 600 μm (A, C, G); 500 μm (E), 300 μm (H), 200 μm (B, D, F).

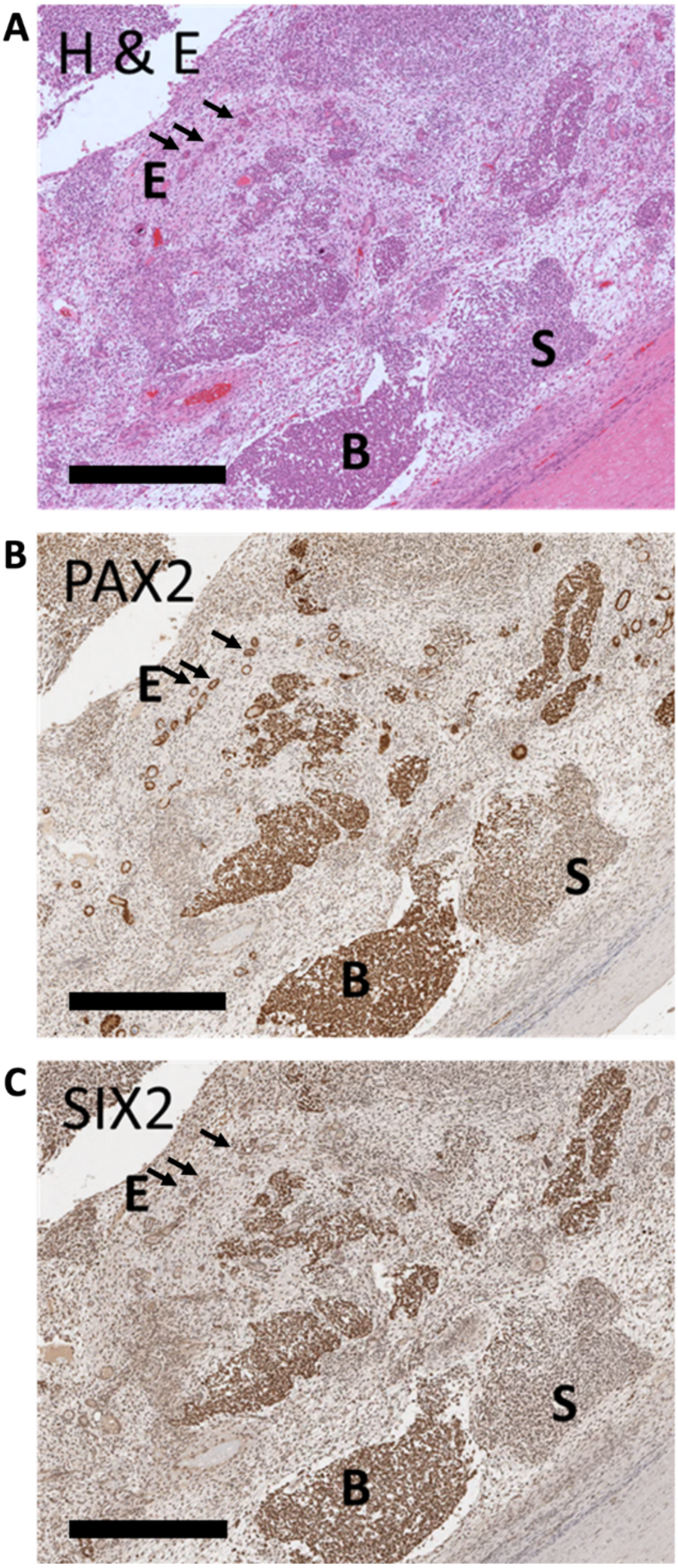


**Supplementary Figure 2. IHC of expression domains of PAX2 and SIX2 in a triphasic WT tissue of index case 1. A.** H&E staining shows epithelial (E), blastemal (B), and stromal (S) components. **B, C.** PAX2 expression is observed in both the blastema and epithelial regions, while SIX2 expression is restricted to the blastema. Scale bars 400 µm.

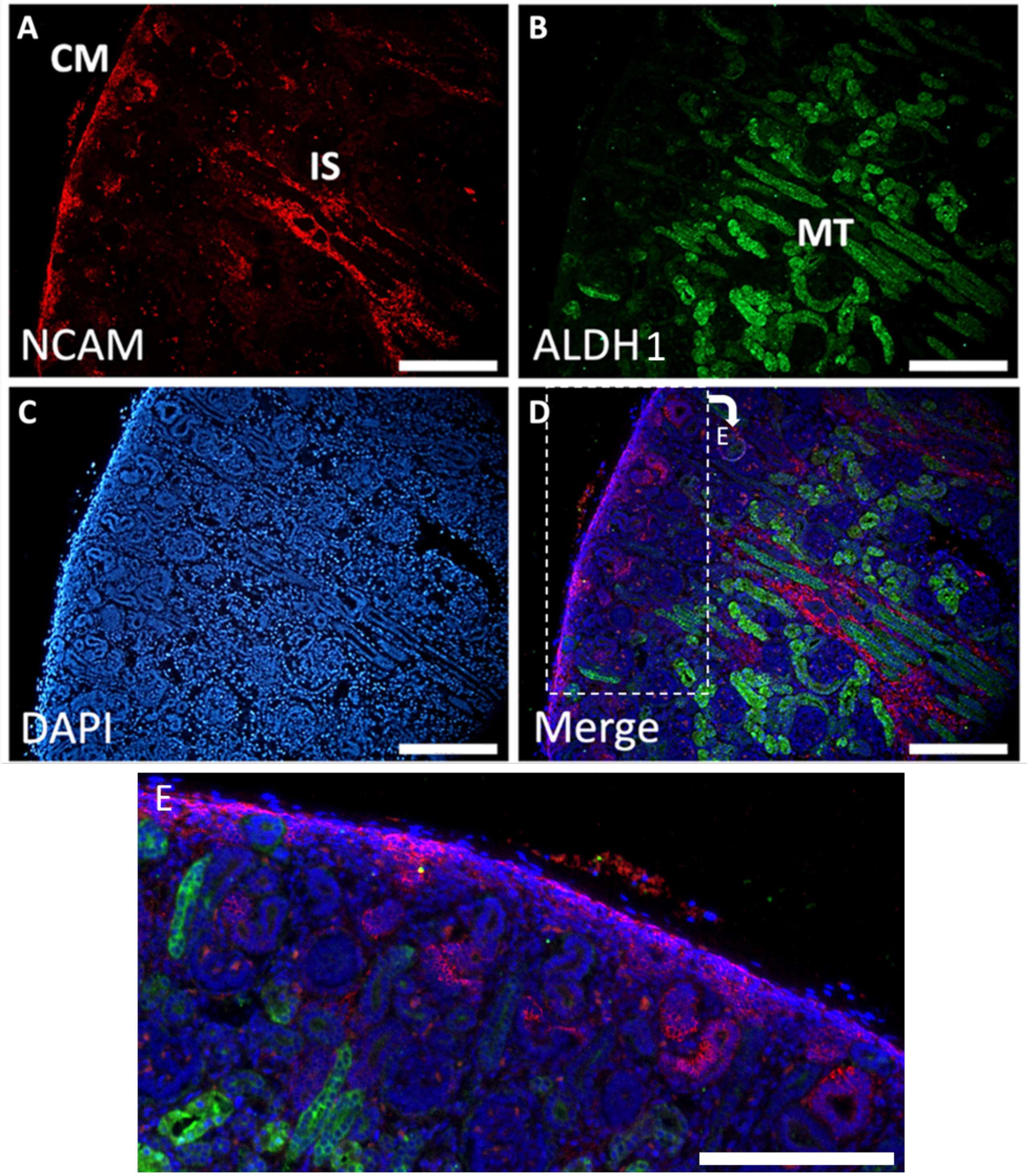


**Supplementary Figure 3. Expression domains of NCAM and ALDH1 in the in hFK.** A-E. IF staining of hFK for NCAM and ALDH1 demonstrated expression of NCAM in the cap mesenchyme (CM) of the nephrogenic zone and in interstitial cells (IS) of the medulla. ALDH1 is detected exclusively in the mature tubule (MT) structures in the medulla. DAPI demarcates nuclei in blue. E is a zoomed-in view of the box outlined in D. Scale bars 100 µm (A-D), 50 µm (E).

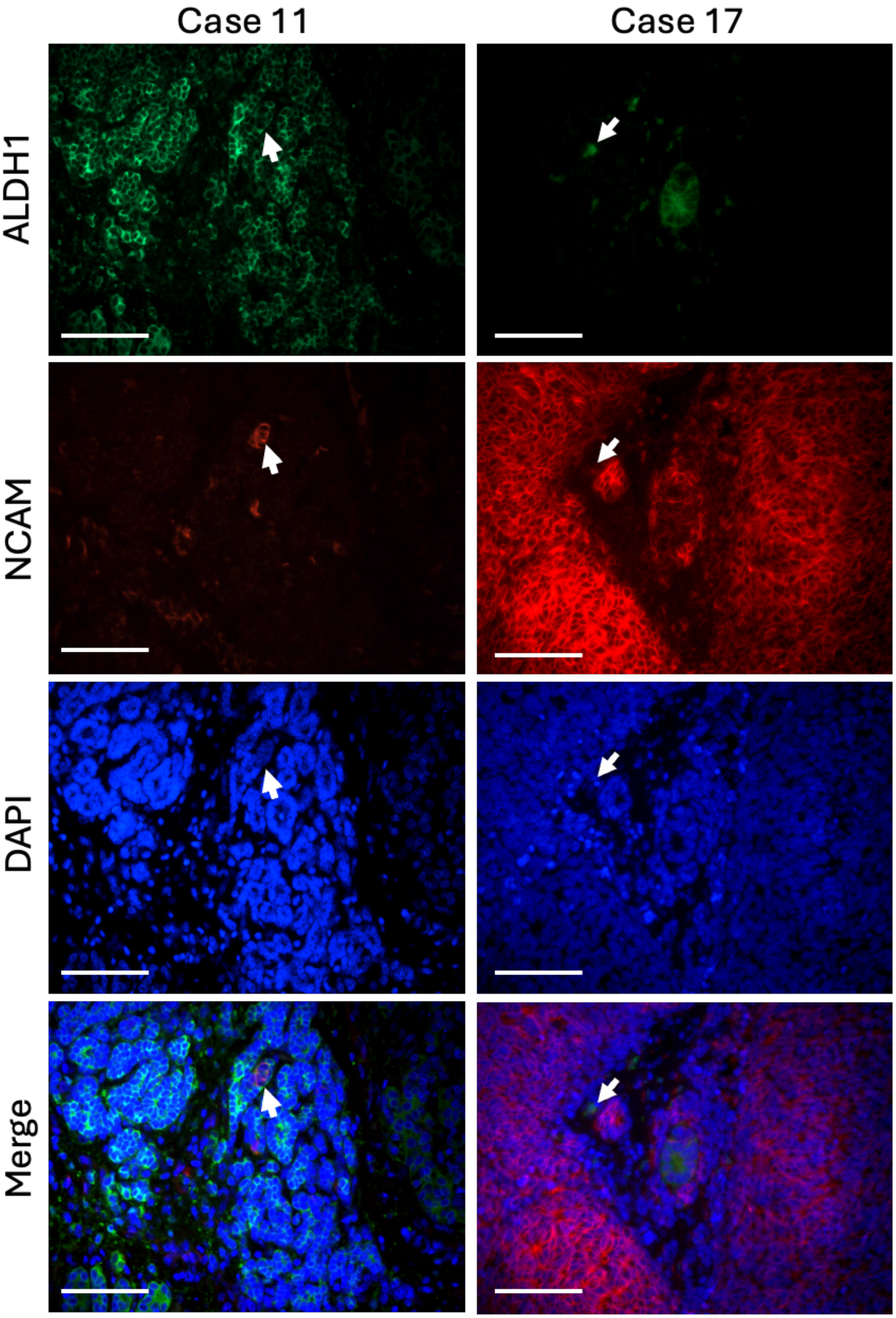


**Supplementary Figure 4. IF double staining of NCAM and ALDH1 in Wilms tumour samples (cases 11 and 17).** The staining demonstrates co-localisation of NCAM and ALDH1 within the blastemal regions in both cases, indicating the presence of NCAM-, ALDH1-co-expressing cells. Scale bars: 200 µm.

A.

| | $PAX2_B$ | $PAX2_E$ | $PAX2_S$ | $SIX2_B$ | $SIX2_E$ | $SIX2_S$ | CITED1 | NCAM | ALDH1 | $NCAM^+$ $ALDH1^+$ | CD133 |
|---|---|---|---|---|---|---|---|---|---|---|---|
| Case 18 | 0 | 1 | 0 | 0 | 0 | 0 | 0 | 0 | 1 | 0 | 0 |
| Case 2 | 1 | 1 | 1 | 0 | 0 | 0 | 1 | 0 | 1 | 0 | 0 |
| Case 1 | 1 | 1 | 0 | 1 | 0 | 0 | 1 | 0 | 1 | 0 | 0 |
| Case 5 | 1 | 1 | 0 | 1 | 0 | 1 | 1 | 1 | 1 | 0 | 0 |
| Case 12 | 1 | 1 | 0 | 1 | 0 | 0 | 1 | 1 | 1 | 0 | 0 |
| Case 15 | 1 | 1 | 0 | 1 | 0 | 0 | 1 | 1 | 1 | 0 | 0 |
| Case 16 | 1 | 1 | 0 | 1 | 0 | 0 | 1 | 1 | 1 | 0 | 0 |
| Case 13 | 1 | 1 | 0 | 1 | 0 | 0 | 1 | 1 | 1 | 0 | 0 |
| Case 3 | 1 | 1 | 0 | 1 | 0 | 0 | 1 | 1 | 1 | 0 | 0 |
| Case 8 | 1 | 1 | 0 | 1 | 0 | 0 | 1 | 1 | 0 | 0 | 0 |
| Case 4 | 1 | 1 | 0 | 1 | 0 | 0 | 1 | 1 | 1 | 1 | 0 |
| Case 11 | 1 | 1 | 0 | 1 | 0 | 0 | 1 | 1 | 1 | 1 | 0 |
| Case 17 | 1 | 1 | 0 | 1 | 0 | 0 | 1 | 1 | 1 | 1 | 1 |
| Case 7 | 1 | 1 | 0 | 1 | 0 | 0 | 1 | 1 | 1 | 1 | 1 |
| Case 10 | 1 | 1 | 0 | 1 | 0 | 0 | 0 | 1 | 1 | 0 | 0 |
| Case 6 | 1 | 1 | 0 | 1 | 1 | 0 | 0 | 1 | 1 | 0 | 0 |
| Case 14 | 1 | 0 | 0 | 1 | 1 | 0 | 1 | 1 | 1 | 0 | 0 |
| Case 9 | 0 | 0 | 0 | 1 | 1 | 0 | 1 | 1 | 1 | 0 | 0 |

B.

| | PAX2 | SIX2 | CITED1 | NCAM | ALDH1 |
|---|---|---|---|---|---|
| Case 1 | 1 | 1 | 1 | 0 | 0 |
| Case 2 | 1 | 0 | 1 | 0 | 0 |
| Case 3 | 1 | 1 | 1 | 1 | 0 |
| Case 4 | 1 | 1 | 1 | 1 | 1 |
| Case 5 | 1 | 1 | 1 | 1 | 0 |
| Case 6 | 1 | 1 | 0 | 1 | 0 |
| Case 7 | 1 | 1 | 1 | 1 | 1 |
| Case 8 | 1 | 1 | 1 | 1 | 0 |
| Case 9 | 0 | 1 | 1 | 1 | 0 |
| Case 10 | 1 | 1 | 0 | 1 | 0 |
| Case 11 | 1 | 1 | 1 | 1 | 1 |
| Case 12 | 1 | 1 | 1 | 1 | 0 |
| Case 13 | 1 | 1 | 1 | 1 | 0 |
| Case 14 | 1 | 1 | 1 | 1 | 0 |
| Case 15 | 1 | 1 | 1 | 1 | 0 |
| Case 16 | 1 | 1 | 1 | 1 | 0 |
| Case 17 | 1 | 1 | 1 | 1 | 1 |
| Case 18 | 0 | 0 | 0 | 0 | 0 |

**Supplementary Figure 5. Clustered heatmaps of co-expression relationships of kidney progenitor and CSC markers across all 18 WT index cases.** Depiction of co-expression of all markers in all tissues (A) or only blastema tissue (B). '1' in blue box indicates expression present, while '0' in grey box indicates no expression. $PAX2_{B/E/S}$, $SIX2_{B/E/S}$ indicate blastemal, epithelial and stromal expression domains for PAX2 or SIX2.
